# Waning of two-dose BNT162b2 and mRNA-1273 vaccine effectiveness against symptomatic SARS-CoV-2 infection is robust to depletion-of-susceptibles bias

**DOI:** 10.1101/2022.06.03.22275958

**Authors:** Kristin L. Andrejko, Jake Pry, Jennifer F. Myers, Megha Mehrotra, Katherine Lamba, Esther Lim, Nozomi Fukui, Jennifer L. DeGuzman, John Openshaw, James Watt, Seema Jain, Joseph A. Lewnard, the California COVID-19 Case-Control Study Team

**Author notes:** Joint corresponding authors: Seema Jain, MD, 850 Marina Bay Parkway, Richmond, California 94804, United States, Kristin Andrejko, PhD, 850 Marina Bay Parkway, Richmond, California 94804, United States. KA and JP contributed equally to the study. Members of the California COVID-19 Case-Control Study Team are listed in the main text.

## Abstract

Concerns about the duration of protection conferred by COVID-19 vaccines have arisen in postlicensure evaluations. However, “depletion of susceptibles” bias driven by differential accrual of infection among vaccinated and unvaccinated individuals may contribute to the appearance of waning vaccine effectiveness (VE) in epidemiologic studies, potentially hindering interpretation of estimates. We enrolled California residents who received molecular SARS-CoV-2 tests in a matched, test-negative design case-control study to estimate VE of mRNA-based COVID-19 vaccines between 23 February and 5 December 2021. We analyzed waning protection following 2 vaccine doses using conditional logistic regression models. Additionally, we used data from case-based surveillance along with estimated case-to-infection ratios from a population-based serological study to quantify the potential contribution of the “depletion-of-susceptibles” bias to time-varying VE estimates for 2 doses. We also estimated VE for 3 doses relative to 0 doses and 2 doses, by time since second dose receipt. Pooled VE of BNT162b2 and mRNA-1273 against symptomatic SARS-CoV-2 infection was 91.3% (95% confidence interval: 83.8-95.4%) at 14 days after second-dose receipt and declined to 50.8% (31.2-75.6%) at 7 months. Accounting for differential depletion-of-susceptibles among vaccinated and unvaccinated individuals, we estimated VE was 53.2% (23.6-71.2%) at 7 months among individuals who had completed the primary series (2 doses). With receipt of a third dose of BN162b2 or mRNA-1273, VE increased to 95.0% (82.8-98.6%), compared with zero doses. These findings confirm that observed waning of protection is not attributable to epidemiologic bias and support ongoing efforts to administer additional vaccine doses to mitigate burden of COVID-19.

## INTRODUCTION

Vaccination has been of critical importance for mitigation of the ongoing COVID-19 pandemic (1). Vaccines currently available in the United States (US) have been found to provide robust protection against severe COVID-19 outcomes including hospitalization and death (2–4). However, suboptimal vaccine uptake in various settings (5), continued public health guidance for isolation of infected and exposed individuals, and lower vaccine effectiveness (VE) among clinically vulnerable patient populations (6–8) highlight the need for robust immunity within the general population to suppress SARS-CoV-2 transmission (9). Post-licensure observational studies (10–12) have estimated declining VE over time following immunization for individuals who received two doses of mRNA-based vaccines. Whereas studies analyzing variant-specific protection have identified only modest differences in VE against Delta (B.1.617.2), Alpha (B.1.1.7), and earlier SARS-CoV-2 lineages among individuals who completed their primary series (13–16), the real-world durability of protection for SARS-CoV-2 infection remains of concern. These considerations have been amplified following the emergence of the Omicron (B.1.1.529) variant, which is associated with additional immune escape (17–19).

In epidemiologic studies, it is critical to distinguish waning of VE from time-varying confounders in the association between vaccination and disease. Under both observational and randomized studies, accrual of immunity through natural exposure to SARS-CoV-2 among unvaccinated persons may gradually erode differences in disease incidence among vaccinated and unvaccinated populations over time (20–23). Termed “depletion-of-susceptibles bias” (24) this phenomenon may merit consideration in studies of COVID-19 vaccines due to the substantial likelihood for cases to evade detection due to asymptomatic or mild clinical presentation (24), particularly in the context of high rates of transmission that have persisted following COVID-19 vaccine rollout in the US. The potential contribution of such biases to reported waning of COVID-19 VE remains unclear (25), along with the role of other factors including patients’ age and clinical risk profile, vaccine product received (26), and spacing of vaccine doses (27).

In California (USA), COVID-19 vaccines became widely available to individuals aged ≥12 years in May 2021, with 68.2% of the eligible population having completed their primary series by 5 December 2021 (28,29). Substantial reductions in COVID-19 cases, hospitalizations, and deaths were observed over the months immediately following COVID-19 vaccine implementation. However, cases, hospitalizations, and deaths partially rebounded from July to October 2021, in association with emergence of the Delta variant as the dominant circulating lineage in the state. Third doses of BNT162b2 were initially recommended on 24 September 2021 for adults aged ≥65 years or who had immunocompromising conditions, who had received BNT162b2 (Pfizer/BioNTech) as their initial vaccine. California subsequently extended eligibility for additional doses of any approved COVID-19 vaccine (mRNA-1273 [Moderna] or Janssen) to all adults aged ≥18 years on 16 November 2021. As of May, 2022, booster doses were recommended for children as young as 5-11 years, and second boosters were available for immunocompromised individuals aged 12 years and older as well as all adults aged ≥50 years.

We analyzed data from an ongoing test-negative design case-control study to characterize differences in VE associated with time since receipt of mRNA-based COVID-19 vaccines, to assess the extent of waning that may be attributable to depletion-of-susceptibles bias, and to determine the effectiveness of administering third mRNA vaccine doses under real-world conditions.

## METHODS

### Recruitment

As part of an ongoing study (29–31), we enrolled California residents with SARS-CoV-2 molecular diagnostic test results reported to the California Department of Public Health (CDPH) between 24 February and 5 December 2021. Analyses were limited to this time window to exclude cases with Omicron variant infection, which accounted for the majority of new-onset SARS-CoV-2 infections in California by late December and is associated with reduced VE (32,33). Trained interviewers administered a questionnaire via phone in English and Spanish each day to individuals whose test result was reported to CDPH in the preceding 48 hours with a recorded telephone number. Cases and control participants were defined as individuals testing positive and negative for SARS-CoV-2, respectively. Individuals were eligible to participate if they had not previously received confirmation of SARS-CoV-2 infection, based on their recollection of any previous positive test result (e.g., molecular, antigen, or serological) or clinical diagnosis. This analysis excludes data from participants aged ≤12 years, who were ineligible to receive vaccination until late in the study period, as well as participants who reported receiving vaccines other than BNT162b2 or mRNA-1273 (National Institutes of Health/Moderna) due to limited observations. The full interview questionnaire is provided in the appendix (**Text S1**).

We sampled cases across nine regions of California with the goal of enrolling participants in equal numbers across each region (**Table S1; Figure S1**). For each enrolled case, interviewers attempted to enroll one control, matched to the case by age category (13-17, 18-29, 30-49, 50-64, 65+ years), sex, and geographic region, from a random sample of individuals testing negative for SARS-CoV-2 whose results were reported to CDPH within the same week. Up to two call attempts were made to enroll cases and controls. Call shifts were scheduled to cover mornings, afternoons, and evenings on both weekdays and weekends to reduce bias in enrollment.

The study protocol was approved as public health surveillance by the State of California Health and Human Services Agency Committee for the Protection of Human Subjects (Project Number: 2021-034).

### Exposures

The primary exposure of interest was each participant’s self-reported COVID-19 vaccination status. Participants who indicated that they had received a COVID-19 vaccine were asked to identify the vaccine manufacturer, number of doses received, and dates of receipt for each dose. Participants were asked to reference their physical vaccination card or another recall aid (e.g., California Digital COVID-19 Vaccine Record at https://myvaccinerecord.cdph.ca.gov, calendar reminder or e-mail confirmation of vaccine appointment, etc.) to confirm their vaccination history during the interview. Participants who completed their primary series of two doses of BNT162b2 or mRNA-1273 more than 14 days before SARS-CoV-2 testing were considered fully vaccinated; other participants who reported receipt of BNT162b2 or mRNA-1273 at the time of testing, and who had not completed their primary series ≥14 days before testing, were considered partially vaccinated. Participants who reported receipt of no COVID-19 vaccine doses at the time of testing were considered unvaccinated. Individuals who reported receiving a third dose prior to their testing date were considered as a separate group.

All participants were further asked to indicate their reasons for seeking a SARS-CoV-2 test and to list all potential symptoms of COVID-19 they had experienced in the 14 days prior to testing; participants who did not indicate experiencing symptoms were prompted with a list of common non-severe symptoms (fever, chills, myalgia, loss of appetite, cough, shortness of breath) to verify their asymptomatic status. We also asked whether participants sought healthcare in association with their SARS-CoV-2 diagnosis including telehealth consultations, outpatient care (at physician offices, urgent care, or retail pharmacy locations), if they presented to an emergency room, and if they were hospitalized. Participants also self-reported whether they had any preexisting or immunocompromising conditions that placed them at higher risk for SARS-CoV-2 infection or COVID-19. We categorized chronic conditions reported by participants using broad classes previously reported by the US Centers for Disease Control and Prevention (34); within our sample, these categories included conditions associated with weakened immune systems, respiratory disorders, cardiovascular or metabolic disorders, and disorders of the liver and/or kidneys (**Table S2**).

### Outcomes

The primary endpoint for our analyses was symptomatic SARS-CoV-2 infection, defined as a positive SARS-CoV-2 test result with ≥1 symptom reported up to 14 days before testing. This symptomatic infection endpoint was the preferred study endpoint because asymptomatic infections were under-represented among cases in this sample, who self-referred for testing; prospective testing strategies would be needed to recruit cases with a representative distribution of infection severity including asymptomatic infections (35,36). As secondary endpoints, we further considered any SARS-CoV-2 infection (regardless of symptoms), infection with fever and ≥1 respiratory symptom reported, and infection for which participants sought or received medical care or advice (beyond testing) in any care setting (virtual, outpatient, emergency department, or inpatient).

### Time-varying protection after second dose receipt

Our primary analysis estimated VE against symptomatic SARS-CoV-2 infection among fully vaccinated participants, in relation to time since receipt of the second dose. Power requirements for determining VE at differing effectiveness thresholds under our study design have been described previously (26). To assess waning of VE, we estimated the adjusted odds ratio (aORs) of prior vaccination among cases versus controls using conditional logistic regression, including an interaction between vaccination status and time from full vaccination (14 days after second dose receipt) to participants’ test date to allow for changes in protection over time. We used the Bayesian Information Criterion (BIC) to compare fit of alternative models formulated with linear, square-root, or logarithmic functions of time since second dose receipt. We defined VE(*t*) = (1 − aOR(*t*)) × 100% to describe the level of protection experienced *t* days after participants were considered fully vaccinated. We defined regression strata according to participants’ age group, sex, week of study enrollment, and geographic region to account for potential confounding in the relationship between vaccination status and infection outcomes.

We repeated these analyses for alternative SARS-CoV-2 infection and symptomatic disease endpoints, redefining the control group as individuals that tested negative for SARS-CoV-2 and met the same thresholds for symptoms as case participants to mitigate potential confounding from association between test-seeking and willingness to be vaccinated (29, 30). We also repeated analyses restricted to participants who reported referencing their vaccination records during telephone interviews to verify robustness of our primary analyses to exposure misclassification.

We undertook the same analyses in subgroups within which we hypothesized that initial VE or risk of waning protection could differ. These included groups defined by product received (BNT162b2 or mRNA-1273), participant age (<50 vs. ≥50 years old), and presence of self-reported preexisting conditions (individuals reporting immunocompetency without any chronic underlying conditions vs. those reporting chronic conditions or weakened immune status). We also undertook subgroup analyses for immunocompetent participants who only reported cardiovascular disease (including hypertension) or obesity as comorbid conditions, as these were the most prevalent conditions reported among all participants.

### Second-dose protection associated with varying inter-dose intervals

Based on previous evidence that spacing between COVID-19 vaccine doses impacts immunogenicity (37), we also assessed whether 2-dose VE and subsequent waning varied in association with the time interval between receipt of the first and second dose (inter-dose interval). We hypothesized that the length of the inter-dose interval could influence the initial strength of protection conferred by two doses, or the persistence of protection over time after receipt of a second dose. We extended the conditional logistic regression frameworks described above to test these hypotheses, assessing improvements in fit of alternative model formulations using BIC. Candidate models included:

1. An interaction between the indicator of having completed the primary vaccination series (receipt of 2 doses >14 days before individuals’ testing date) and days elapsed between first and second doses, defined continuously.
2. An indicator that individuals received their second dose at a longer-than recommended interval (>21 days for BNT162b2 or >28 days for mRNA-1273), interacted with the indicator of having completed the primary vaccination series.
3. An indicator that individuals received their second dose at a longer-than recommended interval, interacted with both the indicator of having completed the primary vaccination series and time since second dose receipt.

### Risk-of-bias analysis

We next sought to determine whether inferences of time-varying VE were robust to depletion-of-susceptibles bias resulting from the differential acquisition rates of natural (infection-derived) immunity among vaccinated and unvaccinated persons within the population (21). While we attempted to reduce such bias in our study design by excluding participants who reported a history of SARS-CoV-2 infection prior to their test, such exclusions are imperfect since a substantial proportion of infections go undiagnosed, especially if symptoms did not occur (38,39). Following previous work (20,22,35), we considered that the aOR of prior vaccination *t* days before testing, comparing cases and controls, would measure the quantity:

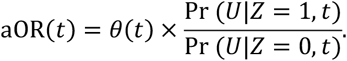

Here we defined *θ*(*t*) as the relative susceptibility of fully vaccinated individuals, as compared to unvaccinated individuals, *t* days after vaccination, owing only to vaccine-derived protection, such that VE(*t*) = (1 − *θ*(*t*)) × 100%. We defined Pr (*U*|*Z, t*) as the probability that individuals remained uninfected (*U*), given their vaccination status (considering *Z* = 1 to indicate fully-vaccinated status and *Z* = 0 to indicate unvaccinated status, for individuals offered vaccination *t* days previously). For simplicity, this formulation did not account for recurrent infections; substantial protection was associated with naturally-acquired immunity for both vaccinated and unvaccinated individuals prior to emergence of the Omicron variant (40), resulting in low risk of re-infection over the time span of several months.

We used the product limit formula to estimate the proportion of fully vaccinated and unvaccinated Californians remaining uninfected by time *t*:

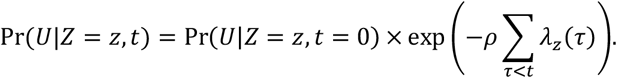

Here, *λ*_*z*_(*τ*) indicated daily incidence rates of SARS-CoV-2 infection among California residents with vaccination status *Z* = *z*, computed as a seven-day moving average around each day *τ* between February and December 2021 (41). To account for the underreporting of cases, the multiplier *ρ* conveyed the ratio of total SARS-CoV-2 infections to reported cases over the period of interest. We used estimates of *ρ* (mean: 2.6, 95% confidence interval: 2.3 to 2.9) from a statewide serological study conducted between April and June 2021 (42), and conducted sensitivity analyses allowing for differing values of *ρ* among vaccinated and unvaccinated persons, which may have arisen due to differences in the likelihood of symptoms or test-seeking between these groups (35). Last, the term Pr(*U*|*Z* = *z, t* = 0) indicated the proportion of individuals who remained uninfected at the time of receiving vaccination, or (for those who remained unvaccinated) at the time vaccination became available to them. We assumed that prevalence of prior infection could differ among vaccine recipients and non-recipients for various reasons, including the initial prioritization of COVID-19 vaccines for populations at high risk of exposure (e.g. essential workers) and conversely, because willingness to receive vaccination could be correlated with other risk-mitigating behaviors. We conducted sensitivity analyses allowing for Pr(*U*|*Z* = *z, t* = 0) equal to 100%, 90%, and 80%, for both the vaccinated and unvaccinated, based on estimates of statewide population seroprevalence as of November 2020 (43).

To estimate bias-corrected VE, we solved for *θ*(*t*) according to for aOR(*t*) estimated via the conditional logistic regression framework described above in our primary analyses.

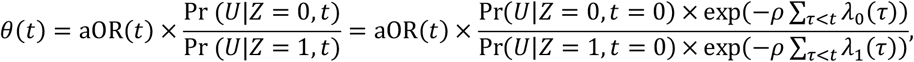

### Protection after third dose receipt

Last, we sought to estimate VE for three doses of BNT162b2 or mRNA-1273 against symptomatic SARS-CoV-2 infection, ≥7 days after receipt of the third dose. We defined three-dose VE relative to receipt of zero doses and two doses at varying time intervals after the second dose (defined categorically by months since individuals were considered fully vaccinated with two doses). Assuming 90% VE of ≥3 doses, we determined that analyses with 85 cases and 85 controls, where 10% of controls had received ≥3 doses, would provide 80% power to infer VE>0% at two-sided *p*<0.05.

We estimated the aOR for prior receipt of three doses, relative to zero doses and two doses, using logistic regression models that included participants’ age group, sex, region, presence of immunocompromising or co-morbid conditions, and the week of study enrollment as categorical covariates. We defined third-dose VE as (1 − aOR) × 100%.

## RESULTS

### Enrollment and descriptive analyses

Between 24 February and 5 December 2021, we enrolled 2238 participants including 1052 cases (testing positive for SARS-CoV-2) and 1186 controls (testing negative for SARS-CoV-2; **Table 1**). In total, 862 participants including 250 cases (24% of 1052) and 612 controls (52% of 1186) reported receiving any mRNA vaccine doses prior to SARS-CoV-2 testing.

**Table 1:**
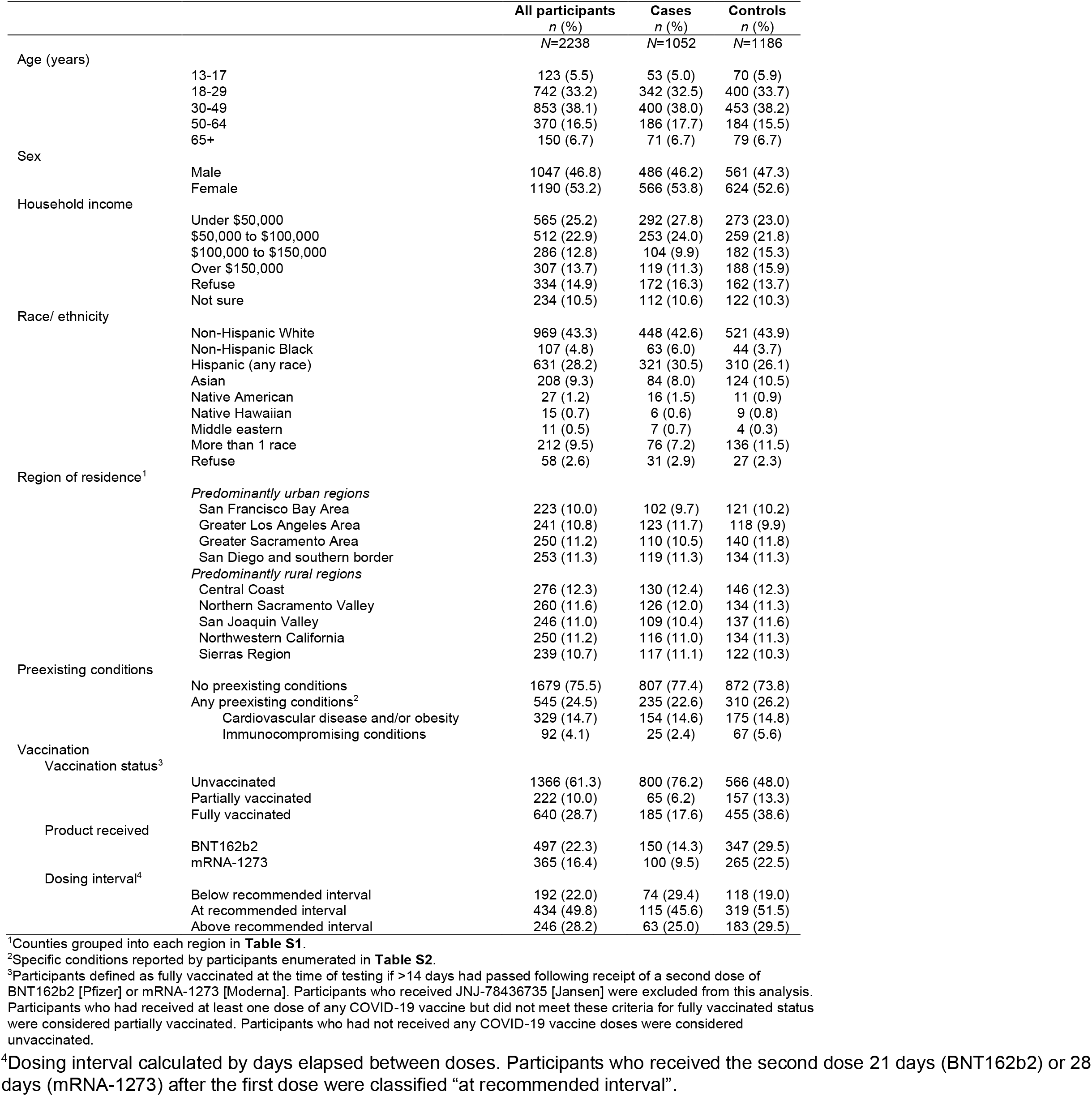
Descriptive attributes of participants included in the analysis of waning vaccine effectiveness against symptomatic infection in California, United States, February to November 2021.

|  |  | All participants<br>*n* (%)<br>*N*=2238 | Cases<br>*n* (%)<br>*N*=1052 | Controls<br>*n* (%)<br>*N*=1186 |
| --- | --- | --- | --- | --- |
| Age (years) | 13-17 | 123 (5.5) | 53 (5.0) | 70 (5.9) |
|  | 18-29 | 742 (33.2) | 342 (32.5) | 400 (33.7) |
|  | 30-49 | 853 (38.1) | 400 (38.0) | 453 (38.2) |
|  | 50-64 | 370 (16.5) | 186 (17.7) | 184 (15.5) |
|  | 65+ | 150 (6.7) | 71 (6.7) | 79 (6.7) |
| Sex | Male | 1047 (46.8) | 486 (46.2) | 561 (47.3) |
|  | Female | 1190 (53.2) | 566 (53.8) | 624 (52.6) |
| Household income | Under \$50,000 | 565 (25.2) | 292 (27.8) | 273 (23.0) |
| | \$50,000 to \$100,000 | 512 (22.9) | 253 (24.0) | 259 (21.8) |
| | \$100,000 to \$150,000 | 286 (12.8) | 104 (9.9) | 182 (15.3) |
| | Over \$150,000 | 307 (13.7) | 119 (11.3) | 188 (15.9) |
|  | Refuse | 334 (14.9) | 172 (16.3) | 162 (13.7) |
|  | Not sure | 234 (10.5) | 112 (10.6) | 122 (10.3) |
| Race/ ethnicity | Non-Hispanic White | 969 (43.3) | 448 (42.6) | 521 (43.9) |
|  | Non-Hispanic Black | 107 (4.8) | 63 (6.0) | 44 (3.7) |
|  | Hispanic (any race) | 631 (28.2) | 321 (30.5) | 310 (26.1) |
|  | Asian | 208 (9.3) | 84 (8.0) | 124 (10.5) |
|  | Native American | 27 (1.2) | 16 (1.5) | 11 (0.9) |
|  | Native Hawaiian | 15 (0.7) | 6 (0.6) | 9 (0.8) |
|  | Middle eastern | 11 (0.5) | 7 (0.7) | 4 (0.3) |
|  | More than 1 race | 212 (9.5) | 76 (7.2) | 136 (11.5) |
|  | Refuse | 58 (2.6) | 31 (2.9) | 27 (2.3) |
| Region of residence<sup>1</sup> | *Predominantly urban regions* |  |  |  |
|  | San Francisco Bay Area | 223 (10.0) | 102 (9.7) | 121 (10.2) |
|  | Greater Los Angeles Area | 241 (10.8) | 123 (11.7) | 118 (9.9) |
|  | Greater Sacramento Area | 250 (11.2) | 110 (10.5) | 140 (11.8) |
|  | San Diego and southern border | 253 (11.3) | 119 (11.3) | 134 (11.3) |
|  | *Predominantly rural regions* |  |  |  |
|  | Central Coast | 276 (12.3) | 130 (12.4) | 146 (12.3) |
|  | Northern Sacramento Valley | 260 (11.6) | 126 (12.0) | 134 (11.3) |
|  | San Joaquin Valley | 246 (11.0) | 109 (10.4) | 137 (11.6) |
|  | Northwestern California | 250 (11.2) | 116 (11.0) | 134 (11.3) |
|  | Sierras Region | 239 (10.7) | 117 (11.1) | 122 (10.3) |
| Preexisting conditions | No preexisting conditions | 1679 (75.5) | 807 (77.4) | 872 (73.8) |
|  | Any preexisting conditions<sup>2</sup> | 545 (24.5) | 235 (22.6) | 310 (26.2) |
|  | Cardiovascular disease and/or obesity | 329 (14.7) | 154 (14.6) | 175 (14.8) |
|  | Immunocompromising conditions | 92 (4.1) | 25 (2.4) | 67 (5.6) |
| Vaccination<br>Vaccination status<sup>3</sup> | Unvaccinated | 1366 (61.3) | 800 (76.2) | 566 (48.0) |
|  | Partially vaccinated | 222 (10.0) | 65 (6.2) | 157 (13.3) |
|  | Fully vaccinated | 640 (28.7) | 185 (17.6) | 455 (38.6) |
| Product received | BNT162b2 | 497 (22.3) | 150 (14.3) | 347 (29.5) |
|  | mRNA-1273 | 365 (16.4) | 100 (9.5) | 265 (22.5) |
| Dosing interval<sup>4</sup> | Below recommended interval | 192 (22.0) | 74 (29.4) | 118 (19.0) |
|  | At recommended interval | 434 (49.8) | 115 (45.6) | 319 (51.5) |
|  | Above recommended interval | 246 (28.2) | 63 (25.0) | 183 (29.5) |

<sup>1</sup>Counties grouped into each region in **Table S1**.
<sup>2</sup>Specific conditions reported by participants enumerated in **Table S2**.
<sup>3</sup>Participants defined as fully vaccinated at the time of testing if >14 days had passed following receipt of a second dose of BNT162b2 [Pfizer] or mRNA-1273 [Moderna]. Participants who received JNJ-78436735 [Jansen] were excluded from this analysis. Participants who had received at least one dose of any COVID-19 vaccine but did not meet these criteria for fully vaccinated status were considered partially vaccinated. Participants who had not received any COVID-19 vaccine doses were considered unvaccinated.
<sup>4</sup>Dosing interval calculated by days elapsed between doses. Participants who received the second dose 21 days (BNT162b2) or 28 days (mRNA-1273) after the first dose were classified "at recommended interval".

Among these 862 vaccinated participants, 497 (58%) and 365 (42%) reported receipt of BNT162b2 and mRNA-1273, respectively. In total, 720 participants were fully vaccinated at the time of their SARS-CoV-2 test. While participants received second doses as few as 14 days after their primary doses, the majority received their second dose at the recommended interval (≥21 for BNT162b2 and ≥28 days for mRNA-1273; **Figure 1A; Figure 1B**). Positive tests among vaccinated cases occurred between zero and 251 days after they were considered fully vaccinated (**Figure 1C**). In contrast, the majority of controls were enrolled within the first 100 days after being considered fully vaccinated (**Figure 1D**).

**Figure 1:**
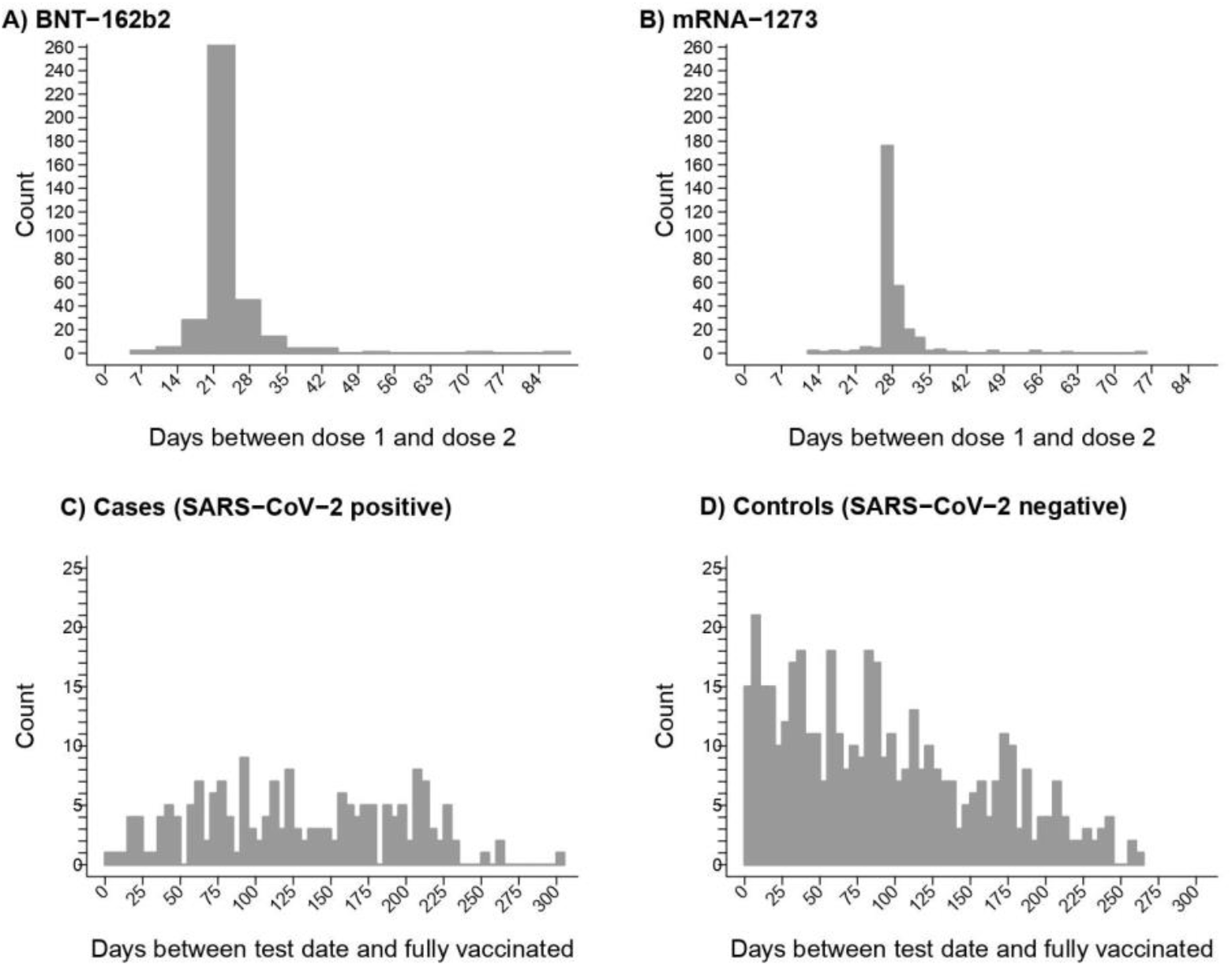
Descriptive attributes of California (USA) residents seeking SARS-CoV-2 infection enrolled between February to December 2021. Distribution of the days elapsed between vaccination and SARS-CoV-2 testing among individuals who received two doses of either BNT162b2 (Pfizer/BioNTech) or mRNA-1273 (NIH/Moderna). Plots show the distribution of BNT162b2 (**A**) and mRNA-1273 (**B**) for which second doses are recommended ≥21 and ≥28 days after first dose, respectively. The distribution shows the days elapsed between participants’ date of full vaccination for cases testing positive for SARS-CoV-2 (**C**) and controls testing negative (**D**). Participants were considered fully vaccinated if they were tested ≥14 days after their second dose of mRNA-1273 or BNT162b2.

### Time-varying protection after second dose receipt

We estimated that the pooled VE for two doses of BNT162b2 or mRNA-1273 against symptomatic SARS-CoV-2 infection was 91.3% (95% confidence interval [CI]: 83.8%, 95.3%) at 14 days after second-dose receipt, but declined thereafter (**Figure 2**). We estimated that VE for two doses of BNT162b2 or mRNA-1273 reached 50.8% (19.7%, 69.8%) seven months after individuals were considered fully vaccinated (absolute difference in VE: 40.2% [17.5%, 73.7%]). Lower bounds of the 95% CI crossed zero at eight months after participants were considered fully vaccinated with two doses (VE=42.9% [–0.1%, 67.1%]; **Table S3**). We obtained similar findings in analyses excluding individuals (*N*=114) who reported receipt of BNT162b2 or mRNA-1273 but did not reference their vaccination record during the interview (**Figure S1**). Estimates were near-identical in analyses restricting the control sample to participants who tested negative for SARS-CoV-2 and reported at least one symptom (**Figure S2B**). Waning was likewise evident in analyses considering endpoints of any SARS-CoV-2 infection (**Figure S2A**), infection with fever and ≥1 respiratory symptom (**Figure S2C**), and infection for which participants received care in any clinical setting (**Figure S2D**).

**Figure 2:**
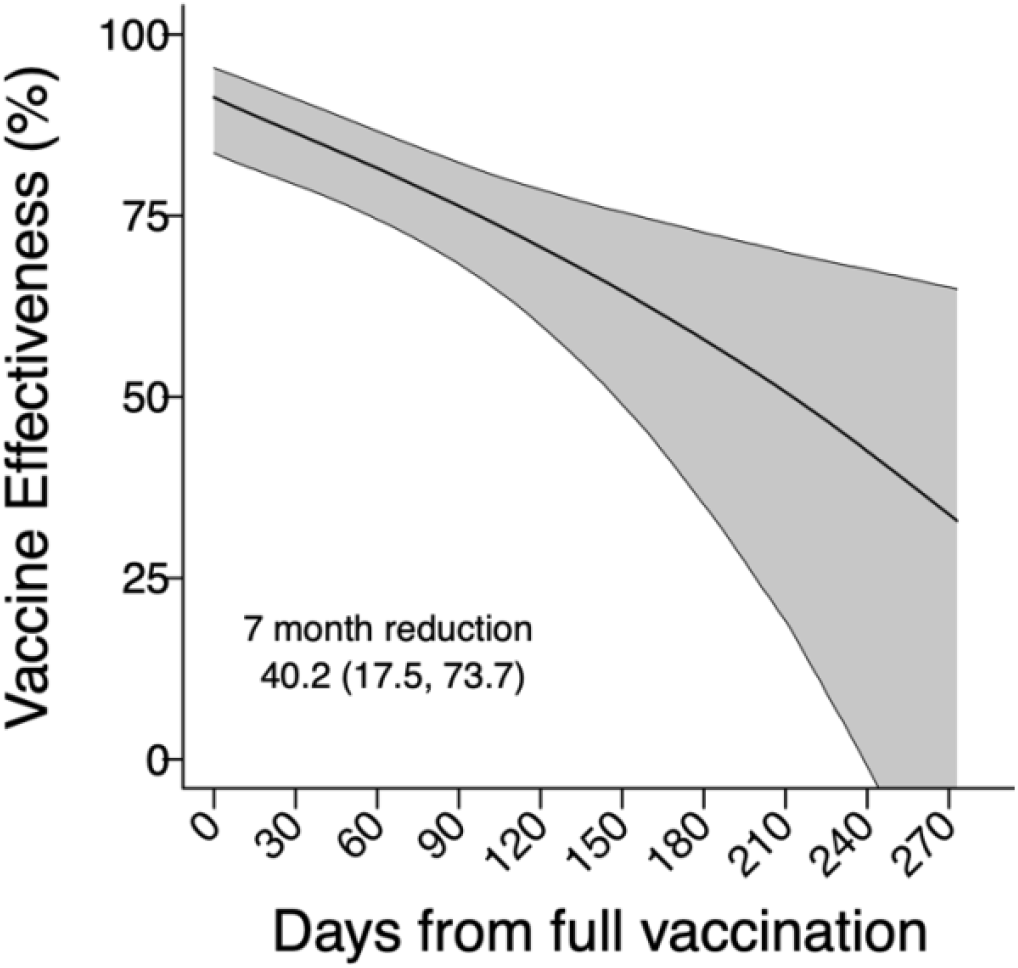
Two-dose vaccine effectiveness against symptomatic SARS-CoV-2 infection over time. Estimates from a conditional logistic regression model matching cases and controls on age group, sex, region, and week of SARS-CoV-2 testing are shown for VE of two doses of BNT162b2 or mRNA-1273 by days since participants were fully vaccinated. Shaded areas denote 95% confidence intervals around point estimates. Numerical estimates plotted in this figure are presented in **Table S3**.

Waning effects did not vary by vaccine manufacturer (**Figure 3A; Figure 3B**). Over a seven-month interval from the date participants were considered fully vaccinated, we estimated 40.5% (17.6%, 73.2%) and 40.4% (17.7%, 74.2%) absolute reductions in VE against symptomatic infection for recipients of BNT162b2 and mRNA-1273, respectively. Over the same interval, participants aged ≥50 years (*N*=520) experienced a 42.6% (10.2%, 103.3%) absolute reduction in VE, as compared to 34.9% (14.4%, 64.0%) absolute reduction in VE among participants aged 13-49 years (*N*=1,718; **Figure 3C; Figure 3D**). Among 545 participants who reported having any preexisting conditions, the reduction in estimated VE over a seven-month interval was 53.0% (–4.1%, 143.4%; **Figure 3F**). However, the estimated reduction in VE was 33.1% (10.3%, 69.1%; **Figure 3E**) among individuals who did not report any preexisting conditions. Among the subset of immunocompetent individuals (*N*=302) who only reported cardiovascular disease (including hypertension) or obesity as comorbid conditions, the seven-month reduction in estimated VE was 37.8% (–36.8%, 149.2%; **Figure 3G**).

**Figure 3:**
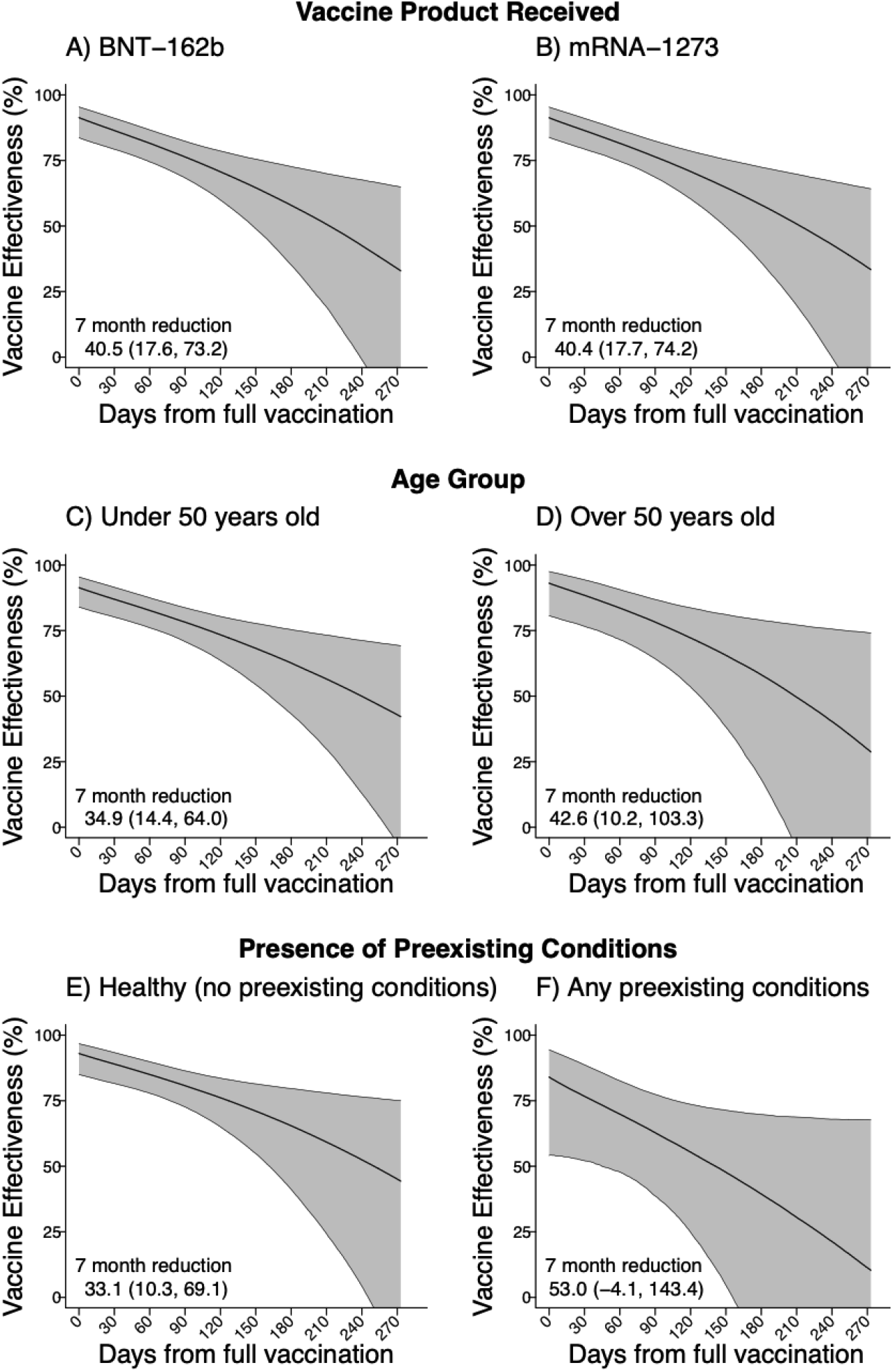
Two-dose vaccine effectiveness against symptomatic SARS-CoV-2 infection over time within distinct participant strata. VE estimates are partitioned for two doses by time since participants were considered fully vaccinated, for: BNT162b2 recipients (**A**), mRNA-1273 recipients (**B**), participants aged 13-49 years (**C**), participants aged ≥50 years (**D**), participants with no self-reported preexisting conditions (**E**); participants who self-reported any preexisting conditions (**F**); and participants who reported cardiovascular disease or obesity, specifically (**G**). Comorbid conditions within the participant sample are enumerated in **Table S2**. Estimates obtained via conditional logistic regression models matching cases and controls on age group, sex, region, and week of SARS-CoV-2 testing. Shaded areas denote 95% confidence intervals around point estimates.

Lengthening the dosing interval from 21 days to 51 days for either BNT162b2 or mRNA-1273 was associated with an absolute increase in VE of 16.3% (–2.4%, 28.7%), at 14 days after receipt of the second dose (**Figure 4A; Figure 4B**). We did not identify evidence for differences in the degree of vaccine waning for participants who received second doses at longer time intervals after their first dose, although these analyses were limited by the low degree of variability in second dose timing (**Figure S3**; **Table S4**).

**Figure 4:**
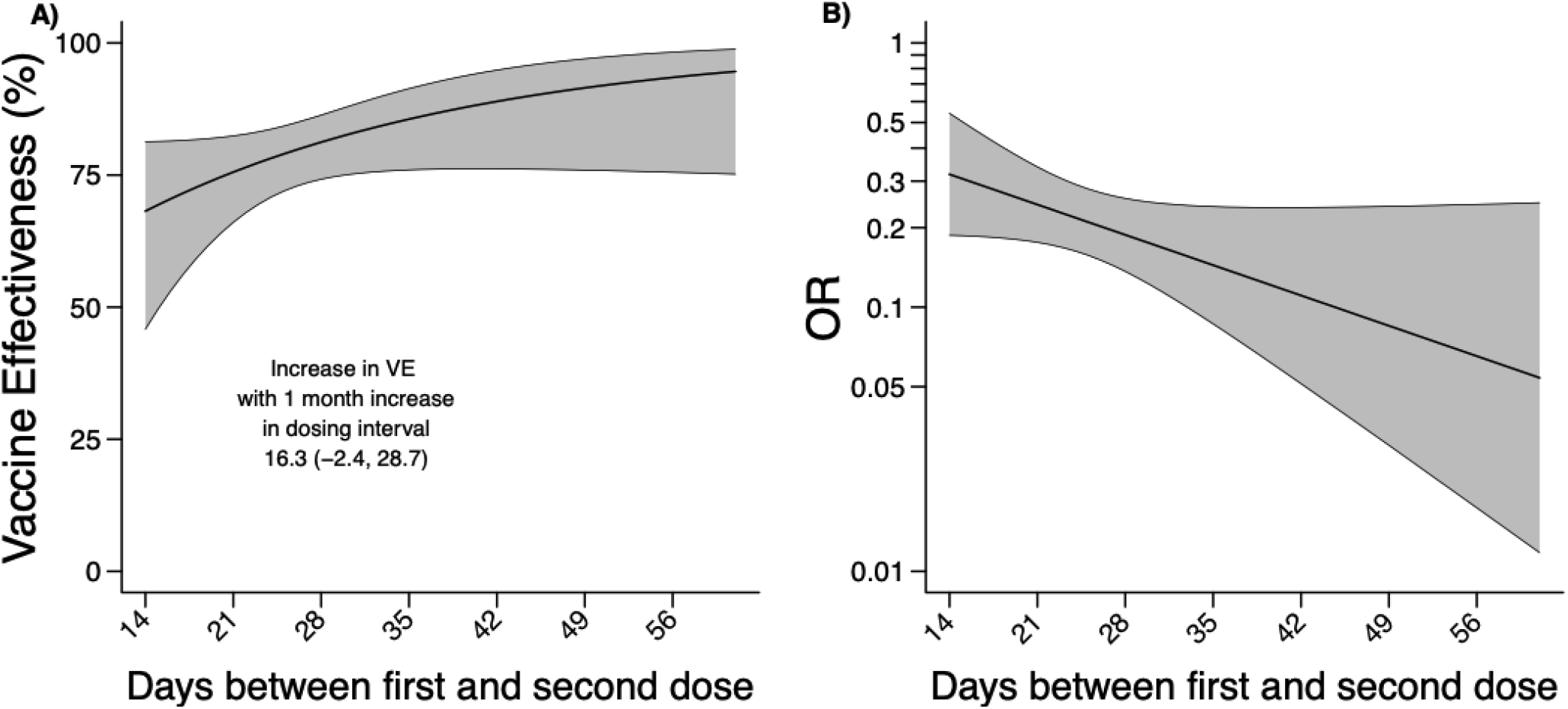
Two-dose vaccine effectiveness against symptomatic SARS-CoV-2 infection by interval between receipt of first and second doses. Two-dose VE estimates presented as of 14 days after second dose receipt, according to the length of the interval between receipt of first and second doses (**A**). The same estimates are presented for the aOR scale (for VE = (1 − aOR) × 100%) to illustrate fold change in susceptibility (**B**). Estimates were obtained via conditional logistic regression models matching cases and controls on age group, sex, region, and week of SARS-CoV-2 testing. Shaded areas denote 95% confidence intervals around point estimates. Longer-term VE for two doses plotted according to the length of the inter-dose interval in **Figure S3**.

### Risk-of-bias analysis

Throughout the study period, incidence rates of reported COVID-19 among unvaccinated persons exceeded those among the vaccinated (**Figure S4**). Incidence rates among the unvaccinated and vaccinated peaked at 90 cases per 100K and 15 cases per 100K respectively, in late July 2021. For a hypothetical cohort that became fully vaccinated as of May 2021, assuming 80% of the population remained uninfected at this time, bias-corrected VE as of December 2021 (7 months after participants were considered fully vaccinated) was 53.2% (23.6, 71.2%), compared to 50.8% (19.7%, 69.8%) without bias correction (**Figure 5**). Estimates of bias-corrected VE did not differ markedly in analyses employing alternative assumptions about the pre-vaccination prevalence of naturally-acquired immunity: sharp bounds on bias-corrected VE ranged from 41.5% (4.5%, 64.1%) when assuming 0% prevalence of prior natural immunity among the unvaccinated to 62.5% (38.9%, 77.0%) when assuming 0% prevalence of prior natural immunity among the vaccinated (**Table S5**).

**Figure 5.**
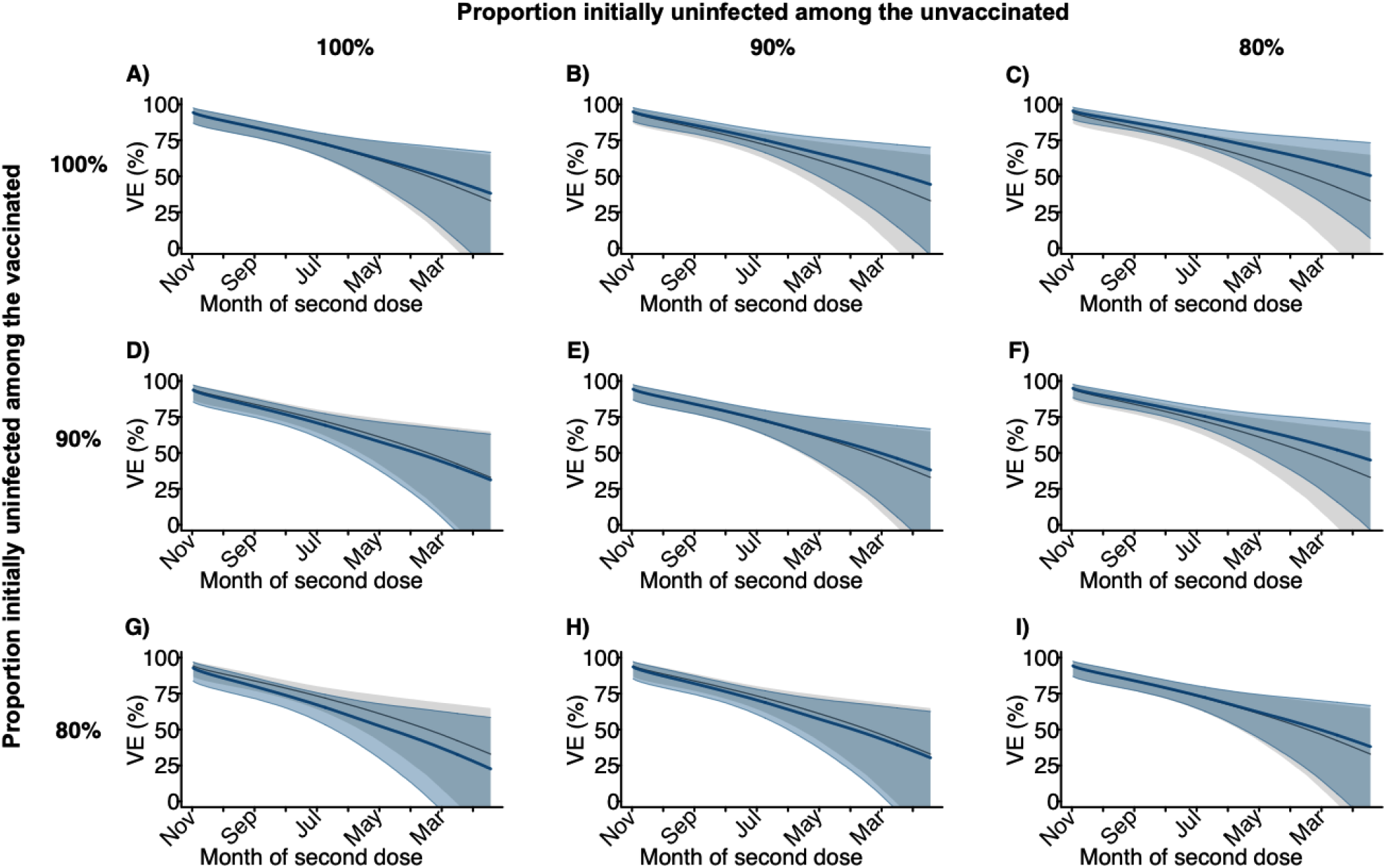
Two-dose vaccine effectiveness against symptomatic SARS-CoV-2 infection as of December 2021 by time since second dose receipt, correcting for depletion-of-susceptibles bias. The “naïve” estimates of two-dose VE from **Figure 2** (grey) were overlayed with bias-corrected estimates (blue) accounting for unreported infections among vaccinated and unvaccinated persons. Rows and columns correspond to differing assumptions about the proportion of individuals who remained uninfected at the time vaccination was offered to them, among the vaccinated and unvaccinated respectively. Scenarios considered include: 100% uninfected among the vaccinated and the unvaccinated, when offered vaccination (**A**); 90% uninfected among the unvaccinated and 100% uninfected among the vaccinated (**B**); 80% uninfected among the unvaccinated and 100% uninfected among the vaccinated (**C**); 100% uninfected among the unvaccinated and 90% uninfected among the vaccinated (**D**); 90% uninfected among the unvaccinated and the vaccinated (**E**); 80% uninfected among the unvaccinated and 90% uninfected among the vaccinated (**F**); 100% uninfected among the vaccinated and 80% uninfected among the unvaccinated (**G**); 90% uninfected among the vaccinated and 80% uninfected among the unvaccinated (**H**); 80% uninfected among the vaccinated and the unvaccinated (**I**). Corresponding numerical estimates are presented in **Table S5**.

### Protection after third dose receipt

We enrolled 3 cases and 22 controls who reported receiving a third mRNA vaccine dose ≥7 days before their SARS-CoV-2 test (**Table 1, Table S6**); these participants received their third dose within 7-76 days before testing. Among all 3-dose recipients, 22 (88.0% of 25) received 3 doses of a single product, while 3 (12.0% of 25) received mixed series. The median time between second and third dose receipt was 213 days. Participants who had received 3 doses of BNT162b2 or mRNA-1273 experienced 95.0% (95%CI: 82.8%, 98.6%) protection against symptomatic SARS-CoV-2 infection relative to unvaccinated participants (**Table 2**). The relative effectiveness of three doses versus two doses 1 month after participants had completed their primary series was 71.6% (–45.4%, 94.3%). The relative effectiveness of three doses versus two doses at 8 months after participants had completed their primary series was 87.9% (51.9%, 97.0%).

**Table 2:** Effectiveness of third doses of BNT162b2 or mRNA-1273 against symptomatic SARS-CoV-2 infection.

| Exposure |  | Cases | Controls | VE of 3 doses, vs. indicated exposure<br>(95% CI) |
| --- | --- | --- | --- | --- |
|  |  | *N* (%) | *N* (%) |  |
| 3 doses |  |  |  |  |
|  | 3 doses ≥7 days before testing | 3 (0.3) | 22 (1.7) | -- |
| 2 doses (fully vaccinated) |  |  |  |  |
|  | Fully vaccinated 1 month prior | 12 (6.5) | 88 (19.3) | 71.6 (–45.4, 94.3) |
|  | Fully vaccinated 2 months prior | 19 (10.3) | 82 (18.0) | 81.1 (11.7,96.0) |
|  | Fully vaccinated 3 months prior | 27 (14.6) | 74 (16.2) | 84.1 (33.6,96.4) |
|  | Fully vaccinated 4 months prior | 28 (15.1) | 56 (12.3) | 85.9 (42.8,96.6) |
|  | Fully vaccinated 5 months prior | 22 (11.9) | 39 (8.6) | 84.4 (37.1,96.3) |
|  | Fully vaccinated 6 months prior | 27 (14.6) | 45 (9.9) | 85.0 (40.5,96.4) |
|  | Fully vaccinated 7 months prior | 24 (13.0) | 28 (6.1) | 85.3 (41.1,96.4) |
|  | Fully vaccinated 8 months prior | 23 (12.4) | 22 (4.8) | 87.9 (51.9,97.0) |
| 0 doses |  |  |  |  |
|  | 0 vaccine doses received at any time | 942 (83.6) | 842 (64.9) | 95.0 (82.8, 98.6) |

## DISCUSSION

Our study demonstrates waning protection against symptomatic SARS-CoV-2 infection with increasing time from receipt of two doses of BNT162b2 or mRNA-1273. The degree of waning VE did not differ between the two vaccines. Depletion-of-susceptibles bias resulting from acquisition of immunity through natural infection did not account for the observed waning of VE with increasing time from receipt of the second dose; estimates correcting for depletion-of-susceptibles bias differed minimally from primary regression-based VE estimates accounting for time-varying protection. These results substantiate previous reports of waning protection up to 5 months after individuals completed their primary vaccination series with BNT162b2 in studies undertaken in the United States (10), Israel (11,44,45), the UK (46), and Qatar (12), demonstrating the persistence of statistically-significant protection against symptomatic infection through 7 months. Encouragingly, administration of a third dose of either BNT162b2 or mRNA-1273 restored protection to levels seen 14 days after second dose receipt (92.4% and 91.3% VE after 3 and 2 doses, respectively), although it should be noted that these findings preceded establishment of the Omicron variant as the dominant circulating SARS-CoV-2 lineage in California. This study’s findings that waning of two-dose VE is not driven by epidemiologic bias, and that VE is restored after a third dose, provide evidence in support of the decision by CDPH, and subsequently CDC, to recommend booster doses for all adults ≥6 months after primary series completion.

Point estimates from our study suggested earlier declines in immunity among older adults (aged ≥50 years) than younger adults (aged <50 years). Although these results should be interpreted cautiously given our limited sample size in older age groups, they are consistent with knowledge that functional changes in innate and adaptive immunity associated with immunosenescence at older ages may reduce the effectiveness of vaccines among older adults (47). Prior studies with greater enrollment of people with older ages have reported similar findings (9, 10). Our analyses also suggested that, as compared to immunocompetent individuals without comorbidities, individuals with weakened immune function or comorbid conditions experienced greater reductions in VE over time after receipt of their second doses (48). It is important to note that our analyses were not powered to compare rates of waning between these populations, and may be subject to misclassification if individuals did not self-report medical conditions accurately or comprehensively; however, these findings likewise resemble those from large-scale studies linking participants’ SARS-CoV-2 testing results to comprehensive medical record datasets (9, 40). Although our analyses address third doses only, our findings are in conceptual agreement with current recommendations prioritizing older and immunocompromised individuals for second booster doses (49).

Extended spacing between the first and second dose of mRNA vaccines was employed in the United Kingdom, Canada, and other countries as a dose-sparing strategy to maximize coverage of first doses. Our findings suggest that longer (>21 or >28 days) intervals between the first and second doses of BNT162b2 or mRNA-1273 may be associated with enhanced clinical protection. In the United Kingdom, individuals receiving second doses 6-12 weeks after their first dose experienced enhanced immunogenicity as well as increased VE against SARS-CoV-2 infection for BNT162b2, and enhanced immunogenicity for ChAdOx1 (Oxford/AstraZeneca) (50). Consideration of extended spacing of doses may thus be appropriate in settings where supply and/or access remains limited.

Our analysis has limitations. The study is observational in nature, and while we attempted to mitigate confounding through matching on test-seeking (by design) as well as age, sex, region, and time, unmeasured confounding in the association between COVID-19 vaccination and individuals’ likelihood of a positive test result may persist. Because we enrolled participants and administered questionnaires via telephone interviews, our study generally did not enroll cases experiencing severe disease. The symptomatic SARS-CoV-2 infection endpoint we monitored should thus be considered to encompass mild or moderate infections, for which most individuals had not been hospitalized by the time of their interviews. Collection of isolates for sequencing was not feasible under our retrospective study design; thus, it was not possible to identify differences in VE across SARS-CoV-2 variants. Data were collected in the study prior to, during, and after the surge of the Delta variant in California, and before emergence of Omicron as a prominent circulating lineage. However, previous analyses distinguishing protection against Delta, Mu, Alpha, and other SARS-CoV-2 variants have suggested that reductions in VE over time are likely due to waning protection over time rather than variant-specific differences in protection (10). Finally, while misclassification of self-reported vaccination status is possible, our VE estimates were robust in analyses excluding participants who did not reference a vaccination card during the interview. Moreover, individuals’ familiarity with referencing their vaccination records to enter restaurants, bars, workplaces, and other public spaces during the COVID-19 pandemic likely reduces the risk for inaccurate reporting of COVID-19 vaccination status relative to other vaccines.

Given the limited duration of follow-up in early COVID-19 vaccine trials, our results demonstrate the value of observational studies to monitor longer-term VE under real-world circumstances. Our findings demonstrate that waning of VE for BNT162b2 and mRNA-1273 is not an artifact of epidemiologic bias, despite such concerns often being raised in studies stratifying VE by age or time since vaccination (17, 18). Our study builds on other findings from the US that third doses of BNT162b2 or mRNA-1273 restore waning VE to levels resembling observations 14 days after receipt of a second dose, thus conferring substantial protection against pre-Omicron variants (51). Similar analyses integrating cumulative infection data from serological studies should be prioritized to distinguish the contributions of naturally-acquired and vaccine-derived immunity to protection against SARS-CoV-2 amid continued circulation of this pathogen, and to characterize the duration of vaccine protection against future SARS-CoV-2 variants.

## Data Availability

Direct data requests to the corresponding author at kristin_andrejko{at}berkeley.edu

## ACKNOWLEDGMENTS

Members of the California COVID-19 Case-Control Study Team include: Adrian Cornejo, Amanda Lam, Amanda Moe, Amandeep Kaur, Anna Fang, Ashly Dyke, Camilla Barbaduomo, Christine Wan, Diana Nicole Morales Felipe, Diana Poindexter, Erin Xavier, Hyemin Park, Helia Samani, Jessica Ni, Julia Cheunkarndee, Mahsa Javadi, Maya Spencer, Michelle Spinosa, Miriam Bermejo, Monique Miller, Najla Dabbagh, Natalie Dassian, Nikolina Walas, Paulina Frost, Savannah Corredor, Shrey Saretha, Sophia Li, Timothy Ho, Vivian Tran, Yang Zhou, Yasmine Abdulrahim, Zheng Dong

## DISCLAIMER

The findings and conclusions in this article are those of the author(s) and do not necessarily represent the views or opinions of the California Department of Public Health or the California Health and Human Services Agency.

## ABBREVIATIONS

CI: Confidence Interval

## SUPPLEMENTAL MATERIAL

### Text S1. Survey Questionnaire

#### SECTION 1 INTRODUCTION (∼2 min)

1. **Hello, my name is [___] and I am calling on behalf of California Department of Public Health to ask some questions regarding [NAME]’s recent COVID-19 test on [INSERT DATE OF TEST]**.
2. *Make sure you’re on the phone with the correct person*. *If case is a child under 18y, make sure you are speaking to a parent/ guardian:*
  2a. **Am I speaking to [NAME]’s parent or guardian?** [If yes, proceed to **<u>section 2</u>**] [If no, proceed to 2b]
  2b. **Can you please pass the phone to [NAME]’s parent or guardian?** [If yes, proceed to **<u>section 2</u>**] [If no, end call] *If case is someone older than 18y:*
  2c. **Am I speaking to [NAME]?** [If yes, proceed to **<u>section 2</u>**] [If no, proceed to 2d]
  2d. **Can you please pass the phone to [NAME]?** [If yes, proceed to **<u>section 2</u>**] [If no- end call]

*NOTE on proxy respondents:*

*If an individual is hospitalized or otherwise too sick to answer questions on their own behalf, a caretaker may serve as a proxy respondent, but verbal consent must first be obtained from the primary case both to participate in the study and to have the proxy respondent answer on their behalf*.

*A proxy respondent who speaks English or Spanish may answer if the individual is unable to easily complete the interview in one of these two languages, provided they are able to speak English or Spanish with sufficient proficiency to provide verbal consent for both participation and for communicating via the proxy respondent*.

#### SECTION 2 ASSENT (∼1 min)

*If you are speaking to a parent or correct person for the first time, add your name and affiliation before starting:*

**Hello, my name is [_____]and I am calling on behalf of California Department of Public Health**.

1. **Hi! We are interested in asking you some questions about [YOUR or INSERT CHILD’S NAME] recent COVID-19 test. We are hoping to interview you to try to better understand the spread of COVID-19. Do you have some time to chat?** INTERVIEWER: pause and wait for person to confirm that they are still on the line, check YES if they say they are willing to chat If they do not have time, select NO
2. **So before we start, I want to make sure you understand that everything I ask you is confidential, protected by California’s strict privacy laws, and is only being used to inform public health. Your answers will not be shared with any other federal, state, or local authorities, and you’re welcome to decline to answer any question. We anticipate this will take about 20 minutes. I know that sounds like a long time, but we really appreciate your time and your answers will help us answer some extremely important questions about COVID-19**. **Do you understand the information I have just shared with you?** INTERVIEWER: check “yes” if the respondent answers yes and if you deem the respondent to be competent to proceed with consent and interviewing; check “no” and thank the respondent for their time if the respondent says no or if you deem the respondent is not competent to proceed with consent and interviewing.] If it seems like the person needs a <u>proxy respondent</u> due to not speaking well enough English or being too sick, you may ask “**Is there anyone who can help you answer my questions?”**. If you get the proxy respondent on the phone, re-introduce yourself by starting at the top of Section 2 with “Hello, my name is…” and add at the end, **“Can you help answer questions on [insert name of case/control’s] behalf?”** NOTE that a proxy respondent must be over the age of 14. *Interviewers then seek consent from the participant, but the question asked will depend on the age of the desired case/control*. [If participant is answering on their own behalf AND they are older then 18] **Great, thank you! To confirm, are you willing to participate in this interview?** [If participant is a child older than 14, answering on their own behalf, first ask for consent from the parent for the child to answer the survey] **Great, thank you! I want to let you know that your child [INSERT CHILD’S NAME] may answer questions on their own behalf. Are you willing to allow [INSERT CHILD’S NAME] to participate in this interview? If not, you can answer questions on their behalf**. *Interviewer: if the child older than 14 joins the call, make sure to reintroduce yourself and explain the purpose of the survey*. [If participant is a child younger than 14, and adult is answering on their behalf] **Great, thank you! Are you willing to answer questions about [INSERT CHILD’S NAME]’s recent exposures as part of this interview?** [If a proxy respondent will answer on behalf of the study participant] **If you are able, I would suggest putting the phone on speakerphone during this interview, so [insert name/ relationship of proxy respondent] can help you**. **[INSERT NAME OF CASE-CONTROL], are you willing to participate in this interview?** **[INSERT NAME OF CASE-CONTROL], do you consent to allow [NAME OF PROXY RESPONDENT] to answer my questions during this interview. Please stay close by [NAME OF PROXY RESPONDENT] in case it is necessary to clarify any points that come up**. [If no or asks to be called back later, proceed to end of the survey] [If consent is provided and case/control is 7-18 years old, proceed to 3] *Interviewer: select the following options based off of the consent pattern:*
  - Participant provided consent on their own behalf
  - Parent provided consent for child <18 yrs
  - Participant provided consent for proxy respondent to answer on their own behalf
  - No consent was provided
3. **No problem. But before we hang up, do you mind quickly sharing why you are unable or unwilling to complete this call?** *Record the free response* [**<u>End call</u>**]
4. **[INSERT CHILD’S NAME] is welcome to stand by or join the call to help answer questions**. [If child joins the call, proceed to 3b, otherwise skip to **<u>section 3</u>**]
  3b. **Hi [INSERT CHILD’s NAME]. My name is [____] and I work with the California Department of Public Health. I’m going to ask you some questions about activities in the past couple of weeks. Are you willing to answer these questions so that we can better understand the spread of COVID-19?** [Proceed to **<u>section 3</u>**]

#### SECTION 3 LAST COVID TEST (∼3 min)

1. **Great, so to start, I want to ask whether you know your COVID-19 test result from [INSERT DATE OF TEST]?** *Record whether they know or don’t know their test result by selecting on of the options:*
  - Subject knows test result and is positive
  - Subject knows test result and is negative
  - Subject does NOT know test result and is positive
  - Subject does NOT know test result and is negative [If yes and they are positive, proceed to **<u>section 4</u>**] [If yes and they are negative, proceed to 3] [If no, and they are negative, proceed to 2] [If no, and they are positive, proceed to 4]
2. **Your COVID-19 test result from [INSERT DATE OF TEST] has come back negative**. *Record one of the following options:*
  - Yes
  - No
  - Don’t know
  - Refuse
  [Proceed to 3]
3. **Have you ever received a *positive* COVID-19 test result or been told by a health care provider that you are positive for COVID-19?** [If no, proceed to **<u>section 4</u>**] [If yes, end-call saying: **Thanks for letting me know. Those are all the questions I have for you. Thank you for your time and I hope you have a nice day**.
4. **Your COVID-19 test result from [INSERT DATE OF TEST] has come back positive. This means you do have coronavirus disease or COVID-19. In my role with CDPH, I cannot provide you with medical advice. If you need any medical information, please call your healthcare provider. One thing I want to be sure of today is that we have a plan for you to follow up with your healthcare provider, so that they can check on any symptoms you may have and assess your risks. Even if you feel okay now, it is important to have someone you can call if you start feeling sick. If you do not have a healthcare provider, you can go to an urgent care facility or the emergency room if you are not getting better or you feel like you are getting worse**. [Proceed to **<u>section 4</u>**] [If the person brings up clinical questions or concerns about their positive test] **Thank you for sharing that concern. In my role with CDPH, I am not able to give you medical advice. I do want to be sure that you get the help you need. If you believe you are having a medical emergency, you should call 911. Some warning signs that you should go to the emergency room for are: trouble breathing, bluish lips or face, pain or pressure in the chest that does not go away, new confusion or trouble waking or staying awake, but there are other symptoms too. Otherwise, you should call your healthcare provider**.

#### SECTION 4 REASONS FOR TESTING (∼3 min)

1. **Next, I’m going to ask you some questions about your COVID-19 test. Can you describe to me why did you choose to get tested on [INSERT DATE OF TEST]?** *Interviewers will select check boxes from the respondent based off of their response, without prompting them from the following list, and will use a write-in option for any additional reasons for seeking testing. After choosing the best answer from the list, confirm your choice the case/control (ex. “So you got tested for pre or post-travel screening?”)*
  - I had contact with someone who tested positive
  - I had contact with someone who had symptoms, but I do not know if they were confirmed to be positive
  - I was told by a public health worker to get tested because I was exposed to a case
  - I was concerned about symptoms I experienced
  - Someone in my household had contact with someone who was positive
  - A person in my household had contact with someone who had symptoms or suspected they had COVID, but we do not know if they are confirmed to be positive.
  - Routine screening for my job
  - Pre or post-travel screening
  - Test required for a medical procedure
  - I just wanted to see if I was infected
  - Don’t know
  - Refuse
  - Other [*interviewer writes in response*]
2. **At the time you were tested on [DATE OF TEST] were you experiencing any COVID-19 symptoms?** *Record Yes/No/Not Sure/ Refuse* [If yes, ask <u>question 4</u>] [If no, proceed to <u>question 5</u>]
3. **Can you please list the symptoms you were experiencing on or 14 days prior to your test on [DATE OF TEST]** *Interviewers will select the symptoms the individuals indicated that they were experiencing. When the respondent is done listing symptoms, the interviewer may prompt, “Are you sure those were all the symptoms you experienced?” and proceed to confirm absence of the 6 most common symptoms (as applicable), in a conversational manner: “No fever, no chills, no muscle pain, no loss of appetite, no shortness of breath, no cough?”* *Select from the following list of symptoms:*
  - Blocked nose
  - Chills
  - Cough
  - Chest pain
  - Diarrhea
  - Muscle pain
  - Fever
  - Headache
  - Hoarseness
  - Loss of appetite
  - Loss of taste
  - Loss of smell
  - Myalgia (muscle pain)
  - Nausea
  - Runny nose
  - Shortness of breath
  - Sneezing
  - Sore throat
  - Stomach pain
  - Sinus pain
  - Sweating
  - Swollen glands
  - Tickle in throat
  - Watery eyes
  - Don’t know
  - Refuse
  - Other
4. **I am now going to read a list of places you may have sought treatment or advice prior to your test on [DATE OF TEST]. After I read the following options, please answer “Yes” or “No”**. *Record Yes/No/ Not Sure/Refuse e for each of the options below*
  - **Did you seek care at an in-person appointment with your usual physician or healthcare provider**
  - **Did you seek care at a telehealth visit or phone appointment with your usual physician or healthcare provider**
  - **Did you seek care at an in-person visit to an urgent care clinic**
  - **Did you seek care at an in-person visit to a healthcare provider at a retail pharmacy**
  - **Did you visit the emergency room?**
  - **Were you admitted to the hospital?**
  - **And just to follow-up, where there any other forms of healthcare from which you sought treatment advice at the time you had your test on [Insert date of test](specify): ______________**
5. **In the 14 days prior to your test (between ADD DATE to ADD DATE) do you know whether you had known or suspected contact with one or more people who may have tested positive for COVID-19?** *Select one of the following options*
  - Yes- contact with one person who was confirmed positive
  - Yes- contact with more than one person who was confirmed positive
  - Yes- contact with one person who I suspected was positive
  - Yes- contact with more than one person who I suspected was positive
  - No known or suspected contact with a positive case
  - Not sure
  - Refuse

[If case indicated they had KNOWN or SUSPECTED Contact, proceed to **<u>section 5, part A</u>**],

[If the case did not have known or suspected contact, proceed to **<u>section 6</u>**]

#### SECTION 5 CONTACT WITH KNOWN OR SUSPECTED CASE (∼8 min)

[If case indicated they had KNOWN or SUSPECTED Contact, proceed to A]

[If case indicated they did NOT have known or suspected contact, proceed to **<u>section 6</u>**]

A. **I’m going to now ask you some questions about the type of contact you had with the person (people) who may have had COVID-19. We are trying to understand sources of exposure and are hopeful that you are willing to answer the questions honestly, knowing that we aren’t looking or expecting any sort of answer**.
  1. **Was the known/ suspected contact someone who lives in your household?** *if plural (contact with* >*1 person*): **Were any of the known/ suspected contact people who lives in your household** *Record Yes, No, Don’t know, Refuse*
  2. **Did the known/ suspected contact occur indoors, outdoors, or both indoors and outdoors?** *if plural (contact with* >*1 person):* **Did the known/suspected contacts occur indoors, outdoors, or both indoors and outdoors?** *Record Indoors, Outdoors, Both indoors and outdoors, Unknown, or Refuse*
  3. **In the 14 days prior to your test (between [ADD 14 DAYS – TEST DATE HERE] to [ADD TEST DATE]), what are the locations where you may have had contact with this person?** *if plural:* **In the 14 days prior to your test (between [ADD 14 DAYS – TEST DATE HERE] to [ADD TEST DATE]), what are the locations where you may have had contact with these people)** *Record the free response answer*
  4. **I am now going to ask you about different precautions you may or may not have been able to take when you came into contact with the known or suspected positive case. Please answer “Yes, No or Not Sure” after each question:** *Record Y/ N/ Not sure for each of the options below:*
    - **Did you come within 6 feet of this person, indoors?** *If plural:* **Did you come within 6 feet of any of these people, indoors?**
    - **Did you come within 6 feet of this person, outdoors?** *If plural:* **Did you come within 6 feet of any of these people, outdoors?**
    - **Did you have physical contact with this person, (ie. handshake, hug)?** *If plural*: **Did you have physical contact with any of these people (ie. handshake, hug)**
  5. **Did you wear a mask the entire time, most of the time, some of the time, or none of the time that you interacted with this person?** *If plural*: **Did you wear a mask the entire time, most of the time, some of the time, or none of the time that you interacted with these people?** *Record which of the statements they agree with from below:*
    - I wore a mask the entire time I interacted with this (these) person(s)
    - I wore a mask most of the time I interacted with this (these) person(s)
    - I wore a mask some of the time I interacted with this (these) person(s)
    - I did not wear a mask during this (these) interaction(s)
    - Not sure
    - Refuse
  6. **Did the person you had known or suspected contact with wear a mask the entire time, most of the time, some of the time, or none of the time when you interacted with them?** *If plural:* **Did the people you had known or suspected contact with wear a mask all, most, some, or none of the time that you interacted with them** *Record which of the statements they agree with from below:*
    - They wore a mask the entire time we interacted
    - They wore a mask most of the time we interacted
    - They wore a mask some of the time we interacted
    - They did not wear a mask during this interaction
    - Not sure
    - Refuse
  7. **Did you spend more than 3 consecutive hours with this person in the 14 days prior to your test (between Date to Date)**. *If plural*: **Did you spend more than three consecutive hours with these people in the 14 days prior to your test (between Date to Date)** *Record Yes/ No/ Don’t know/ Refuse*

[proceed to **<u>section 6</u>**]

#### SECTION 6 EXPOSURE WITH CONTACT KNOWN OR SUSPECTED CASE (∼10 min)

**Next, I want to learn about potential sources of exposure to COVID-19 in the 14 days before your last test: from [ADD 14 DAYS – TEST DATE HERE] to [ADD TEST DATE]. It may help you to pull up a calendar to remember what you were up to over the last two weeks. This chunk usually takes the longest, so thank you in advance for your time**

*Only read the following if they did <u>not</u> have known or suspected contact:*

**[We are trying to understand sources of exposure and are hopeful that you are willing to answer the questions honestly, knowing that we aren’t looking or expecting any sort of answer.]**

1. **I am now going to ask you about a series of locations which you may have visited. After I announce each location, please tell me “Yes, No, or Not sure” to indicate whether you visited that location between [ADD 14 DAYS – TEST DATE HERE] to [ADD TEST DATE]**.
  - **First, did you attend a health appointment or health facility (other than where you got tested for COVID-19)**
  - **Did you go grocery shopping?**
  - **Now I am going to ask you about the times you went to restaurants. Did you go to any restaurants to pick up take-out or to eat at the restaurant?** *Record one of the following options: a) Dine-in (eat at restaurant) only, b) Take-out only, c) Both dining-in and take-out, d) Neither dine-in or take-out, e) Not sure, f) Refuse* *If yes and take-out:*
  - **How many times did you get take-out?**
  - **Did you ever have to go inside the restaurant to either place or pick up your take-out order?** *Record one of the following options: a) Yes, I went inside the restaurant either to place or pick-up my order, b) No I did not go inside the restaurant either to place or pick-up my order, d) No I did not go inside the restaurant either to place or pick-up my order, but someone I went to the restaurant with had to go inside to place or pick-up the order, e) not sure, f) refuse* *If yes and dine-in:*
  - **How many times did you eat at an <u>indoor</u> restaurant?**
  - **How many times did you eat at an <u>outdoor</u> restaurant?** [Skip the following question chunk about bars if respondent is under 21]
  - **Did you attend any bars, breweries, or wine bars?** *If yes, ask:* **Did you attend a bar, brewery or wine bar?** *Select all For each of the places they indicated that they visited:*
  - **How many times did you attend a [bar/brewery/wine bar]?**
  - **When you went to a (those) [bar(s)/brewery(ies)/wine bar(s)], did you spend most of your time indoors, outdoors, or both indoors and outdoors?**
  - **Did you ever visit a coffee shop?** *If yes, ask:*
  - **When you (typically) visited the coffee shop(s), did you have to go inside to place your order?** *Record one of the following options: a) I went inside to place the order, b) I typically placed the order outside or remotely (via. App, web portal, phone order), c) Don’t know, d) refuse*
  - **When you visited a coffee shop, did you (typically) consume your beverage inside the shop, outside the shop, or did you just pick-up the beverage for take-away**. *Record one of the following options a) consumed inside the shop, b) consumed outside the shop (ex. restaurant set up outdoor tables/ chairs and I drank/ate at those tables), c) Got beverage for take-away, d) Don’t know, e) refuse*
  - **Did you go retail shopping?** *If yes, ask:* **And did you go indoor or outdoor retail shopping?**
  - **Did you exercise at gym?** *If yes, ask:* **And was this an indoor or an outdoor gym?**
  - **Did you participate in a group recreational sport (tennis, soccer, basketball, swimming)**
  - **Did you ever leave your house to go for a walk, run, hike or ride a bike outside?** *If yes ask:* **Did you hike, run, walk, or bike with anyone outside your household?** *Select one of the following options a) No, I always hiked, ran, walked, or biked by myself, b) No, but I sometimes/ always ran, walked, or biked with other people who live in my household, c) Yes I hiked ran, walked, or biked with someone who doesn’t live in my household, d) Don’t know, e) Refuse*
  - **Did you ride public transit?**
  - **Did you use a ride share (eg. Taxi, Uber, Lyft, or carpool with individuals who are not members of your household) ?**
  - **Did you fly on a plane?**
  - **Did you attend a parade, rally, march, or protest?**
  - **Did you receive services at a salon or barber?**
  - **Did you attend an indoor movie theater?**
  - **Did you attend a worship service?** *If yes ask:* **And was this an indoor or an outdoor worship service?**
  - **Did you visit or stay at a school, daycare or preschool?** *If yes, ask:* **Was the school or daycare public or private?**
  - **Did you visit a jail, prison, or correctional facility?**
  *If a participant answers yes to any of the questions in 1, follow-up with:* **How many times did you attend [INSERT LOCATION] between [ADD 14 DAYS – TEST DATE HERE] to [ADD TEST DATE]**. **I am now going to ask you (a couple more) some questions about face mask usage between date to date**.
2. **Between [ADD 14 DAYS – TEST DATE HERE], at all of the <u>indoor</u> places we discussed earlier, did you wear a face mask all, most, some, or none of the time?**
  - I wore a face mask all of the time
  - I wore a face mask most of the time
  - I wore a face mask some of the time
  - I never wore a face mask in indoor places
  - I did not go inside any indoor places other than my home
  - I was not in contact with anyone
3. **Between [ADD 14 DAYS – TEST DATE HERE], at all of the <u>indoor</u> places we discussed earlier, did people you came within 6 feet of wear a face mask all, most, some, or none of the time?**
  - They wore a face mask all of the time
  - They wore a face mask most of the time
  - They wore a face mask some of the time
  - They never wore a face mask in indoor places
  - I did not go inside any indoor places other than my home
  - I was not in contact with any people outside my household in indoor places
4. **Between [ADD 14 DAYS – TEST DATE HERE], at all of the <u>outdoor</u> places we discussed earlier, did you wear a face mask all, most, some, or none of the time?**
  - I wore a face mask all of the time
  - I wore a face mask most of the time
  - I wore a face mask some of the time
  - I never wore a face mask in indoor places
  - I did not go inside any outdoor places other than my home
5. **Between [ADD 14 DAYS – TEST DATE HERE], at all of the <u>outdoor</u> places we discussed earlier, did people you came within 6 feet of wear a face mask all, most, some, or none of the time?**
  - They wore a face mask all of the time
  - They wore a face mask most of the time
  - They wore a face mask some of the time
  - They never wore a face mask in indoor places
  - I did not go inside any outdoor places other than my home
  - I was not in contact with any people outside my household in outdoor places
6. **I am now going to ask you some questions about social gatherings. These include any informal gatherings with friends or family who are NOT members of your household). Did you attend any social gatherings between (14 days prior to test result to test result date)?** *Interviewer: note that our definition of social gatherings is mixing with people who don’t otherwise live in your household. If someone had a longer-term family* together *(ie. traveled to visit relatives, but stayed for multiple days, count this as ONE event)*. *If yes, ask:* **When you attended social gatherings, were they indoors, outdoors, or both indoors and outdoors?** *An <u>outdoor only</u> gathering means the person spent the majority of their time outside An <u>indoor only</u> gathering means the person spent the majority of their time* *A gathering that was “<u>both indoors and outdoors</u>” means the participant was both inside and outside during the social gathering (ex. Sandy had some friends over for dinner and they ate outside on the patio, and then watched a movie in their living room together)* *If they indicate they attended indoor social gatherings***: How many indoor social gatherings did you attend between (14 days prior to test result to test result date)? About how many people attended these gatherings? Did you eat or drink during any of these (or this) gatherings? When you attended this (these) indoor gathering(s), did you wear a face mask all, most, some or none of the time?** *Interviewer: note that this question about mask usage is distinct from the question earlier*. *If they indicate they attended outdoor social gatherings:* **How many outdoor social gatherings did you attend between (14 days prior to test result to test result date)? About how many people attended these gatherings? Did you eat or drink during any of these (or this) gatherings? When you attended this (these) outdoor gathering(s), did you wear a face mask all, most, some or none of the time?** *If they indicate they attended social gatherings that were both inside and outside:* **How many social gatherings did you attend between (14 days prior to test result to test result date) that were both indoor and outdoor? About how many people attended these gatherings? Did you eat or drink during any of these (or this) gatherings? When you attended this (these) outdoor gathering(s), did you wear a face mask all, most, some or none of the time?**
  - **Did you attend any other kind of event where there are <u>5 or more</u> people who are not in your household in attendance?** *Interviewer: If necessary, prompt with options like a sporting event, concert, festival, etc*. **Specify the event: _______________**

[Proceed to **<u>section 7</u>**]

#### SECTION 7 OCCUPATION (∼1 min)

1 **I am now going to ask you some questions about your occupation. Between [ADD 14 DAYS – TEST DATE HERE] to [ADD TEST DATE] did you attend work, school, or volunteering commitments exclusively at home, both at home and in “in-person”, or exclusively “in-person”**.
  ∘ I work, study, and/or volunteer at home
  ∘ I attend work, school, and/or volunteering “in-person”
  ∘ I attend work, school, and/or volunteering both “in-person” and at home
  ∘ I am not currently working, in school, or in a volunteer position.
  [If respondent is a student, skip question and just record “student”]
2 **Can you tell me what your job is?** (*Record open ended response]* [If they attend work, school, or volunteering commitments in person or both at home & in person, proceed to question 3, otherwise proceed to **<u>Section 8</u>**]
  3a. **Do you come into close contact (within 6 feet) of more than 10 people per day at work/school/volunteering?** *Record: Yes or No*
  3b. **Do you primarily attend work/school/volunteering indoors, outdoor, or both indoors and outdoors?** *Record: indoors, outdoors, or both*

[Proceed to **<u>section 8</u>**]

#### SECTION 8 VACCINATION (∼2 min)

**I am now going to ask you some questions about the COVID-19 vaccine**.

1. **Do you have any conditions that might place you higher risk for COVID-19?** *Interviewers may prompt with examples such diabetes, high blood pressure, overweight, being immunocompromised if requested. Select options from list below*
  - Lung conditions: COPD, lung cancer, cystic fibrosis, moderate to severe asthma, pulmonary fibrosis
  - Heart disease
  - High blood pressure
  - Obesity
  - Overweight
  - Diabetes
  - Weakened immune system: organ transplant, cancer treatment, bone marrow transplant, HIV/AIDS, sickle cell anemia, thalassemia
  - Chronic kidney disease
  - Chronic liver disease
  - Pregnant (first, second, or third trimester)
2. **Have you received any doses of a COVID-19 vaccine?** *Record: Yes, No* [If they have not received any doses, skip to 2b, otherwise ask question 3]
  2b. **Do you plan to receive any doses of the COVID-19 vaccine?** *Record: Yes, No, not sure, refuse* [If they are not planning to receive any doses or are not sure yet, ask 2c, otherwise, ask skip to **<u>section 9</u>**]
  2c. **Can you describe to me why you are not planning to receive the COVID-19 vaccine?** *Record reason in check box*
3. **How many doses of the COVID-19 vaccine have you received?** *Record: 1, 2*
4. **Do you have a vaccine card on hand from when you got the COVID-19 vaccine?** *If yes, ask them to get their vaccine card. If no, ask them to do their best remembering and try pulling up a calendar to help them remember*.
5. **What dates did you receive your dose(s)?** *Record the date of each vaccine*
6. **Do you know what product COVID-19 vaccine you received?** *Record the product of each dose*
7. **Do you have access to a COVID-19 vaccination clinic at your work or school?** *Record: yes/ no/ not sure/ refuse*
8. **Where did you get your COVID-19 vaccine?** *Record: mass vaccination site, hospital, nursing home, at my work, at my school, at a retail pharmacy, at a retail shop (eg. Walmart)*
9. **At the time you received the vaccine was it required to attend work or school?** *Record: yes/ no/ not sure/ refuse*

[Proceed to **<u>section 9</u>]**

#### SECTION 9 DEMOGRAPHICS (∼5 min)

**I just have a few more questions. Again, anything you share with me is confidential and protected by California’s strict privacy laws. The information we collect about you will assist the health department in their COVID-19 response**.

1. **First, I’m going to ask you some general questions about COVID-19. From the beginning of the pandemic to the time you were tested on [DATE OF TEST], how worried did you feel about getting COVID-19? Would you say you felt:**
  - **Very worried**
  - **Somewhat worried**
  - **Neutral**
  - **Not worried at all**
2. **Since the beginning of the pandemic, there have been a lot of recommendations on behaviors that can reduce the risk of COVID-19 including avoiding large crowds, travel, and maintaining 6 feet of distance in public places. Would you say that you strongly agree, agree, are neutral, disagree, or strongly disagree that these measures reduce the risk of COVID-19?** *Record strongly agree, agree, neutral, disagree, strongly disagree*
3. **Another recommendation to reduce the spread of COVID-19 is wearing face masks. Would you say that you strongly agree, agree, are neutral, disagree, or strongly disagree that face masks reduce the risk of COVID-19?** *Record strongly agree, agree, neutral, disagree, strongly disagree* **Last, I want to capture some information about demographics**.
  1. **So do you mind sharing how old you are?** *Record free response*
  2. **Next, please let me know which of the following race/ethnicities best describe yourself. You may select all that apply:**
    - **White**
    - **Black**
    - **Hispanic**
    - **Asian**
    - **Native American or Alaska Native**
    - **Native Hawaiian or other Pacific Islander**
  3. **What is your sex/ gender?** *Record Man, Woman, Non=-binary, Prefer to self-describe, Refuse, Don’t know*
  4. **What is your zip code of your home address?** *Record address using encryption tool*
  5. **What is your home address?** *Record address using encryption tool after verifying it is an address using google maps*
  6. **Which of the following best describes your living arrangement:**
    - **Private home**
    - **Apartment, or condominium**
    - **Skilled nursing facility**
    - **College or university student housing**
    - **Military quarters**
    - **Emergency or transitional shelter**
    - **Other (please describe)**
  7. **How many people live in your household?**
  8. **How many bedrooms do you have in your household?**
  9. **Do you have any children under 18 at your home?**
  10. **Are any of your children under 18 attending in-person instruction, school, or daycare?**
  11. **Does anyone visit your home on a regular basis like a cleaning service or babysitter?** If you are talking to a child aged 14-17, at this point you can end the interview with the child and ask to speak with their parent/guardian. When you get back on the phone with the parent or guardian, you can say something like **[“Hi again, thank you so much for letting me speak with your child, it was extremely helpful. We are wrapping up the survey with some demographic questions and my last question that I didn’t want your child to have to answer was whether you are willing to share your total household income?”]**
  12. **What is your total household income? Answer on behalf of everyone you share finances with**. [If you are speaking with POSITIVE case, proceed to 13] [If you are speaking with NEGATIVE control, proceed to 14]
  13. **Thank you for participating in this survey. You may be contacted by another staff member at the health department to check in on you. They will ask you questions about your health and well-being to make sure you’re ok**.
  14. **Thank you for participating in our survey. We appreciate your time**.

**Table S1:** Counties included in each geographic region.

| County | Region |
| --- | --- |
| Alameda County | San Francisco San Francisco Bay Area |
| Alpine County | Sierras Region |
| Amador County | Sierras Region |
| Butte County | Northern Sacramento Valley |
| Calaveras County | Sierras Region |
| Colusa County | Northern Sacramento Valley |
| Contra Costa County | San Francisco Bay Area |
| Del Norte County | Northwestern California |
| El Dorado County | Sierras Region |
| Fresno County | San Joaquin Valley |
| Glenn County | Northern Sacramento Valley |
| Humboldt County | Northwestern California |
| Imperial County | San Diego and southern border |
| Inyo County | Sierras Region |
| Kern County | San Joaquin Valley |
| Kings County | San Joaquin Valley |
| Lake County | Northwestern California |
| Lassen County | Sierras Region |
| Los Angeles County | Greater Los Angeles area |
| Madera County | San Joaquin Valley |
| Marin County | San Francisco Bay Area |
| Mariposa County | Sierras Region |
| Mendocino County | Northwestern California |
| Merced County | San Joaquin Valley |
| Modoc County | Sierras Region |
| Mono County | Sierras Region |
| Monterey County | Central Coast |
| Napa County | San Francisco Bay Area |
| Nevada County | Sierras Region |
| Orange County | Greater Los Angeles area |
| Placer County | Sierras Region |
| Plumas County | Sierras Region |
| Riverside County | Greater Los Angeles area |
| Sacramento County | Central Valley |
| San Benito County | San Francisco Bay Area |
| San Bernardino County | Greater Los Angeles area |
| San Diego County | San Diego and southern border |
| San Francisco County | San Francisco Bay Area |
| San Joaquin County | San Joaquin Valley |
| San Luis Obispo County | Central Coast |
| San Mateo County | San Francisco Bay Area |
| Santa Barbara County | Central Coast |
| Santa Clara County | San Francisco Bay Area |
| Santa Cruz County | San Francisco Bay Area |
| Shasta County | Northwestern California |
| Sierra County | Sierras Region |
| Siskiyou County | Northwestern California |
| Solano County | San Francisco Bay Area |
| Sonoma County | San Francisco Bay Area |
| Stanislaus County | San Joaquin Valley |
| Sutter County | Northern Sacramento Valley |
| Tehama County | Northern Sacramento Valley |
| Trinity County | Northwestern California |
| Tulare County | San Joaquin Valley |
| Tuolumne County | Sierras Region |
| Ventura County | Greater Los Angeles area |
| Yolo County | Northern Sacramento Valley |
| Yuba County | Northern Sacramento Valley |

**Table S2:** Conditions included in each co-morbidity group.

| Condition<sup>1</sup> | Description | Total<br>N=534<br>N (%) | Cases<br>N=234<br>N (%) | Controls<br>N=300<br>N (%) |
| --- | --- | --- | --- | --- |
| Chronic lung conditions | Asthma, Chronic obstructive pulmonary disease (CDPH, Cystic Fibrosis, Pulmonary Fibrosis, lung cancer, bronchiectasis, pulmonary embolism, pulmonary hypertension, bronchopulmonary dysplasia, | 172 (32.2) | 75 (32.1) | 97 (32.3) |
| Immunocompromising conditions | Organ transplant, bone marrow transplant, HIV/AIDS, Cancer Treatment, Rheumatoid arthritis | 87 (16.3) | 24 (10.3) | 63 (21.0) |
| Obesity | Overweight, Obese, Diabetes | 94 (17.6) | 49 (20.9) | 45 (15.0) |
| Cardiovascular disease | Heart disease, hypertension, heart failure, coronary artery disease, cardiomyopathies | 259 (48.5) | 122 (52.1) | 137 (45.7) |
| Liver conditions | Chronic liver disease | 3 (0.6) | 2 (0.9) | 1 (0.3) |
| Kidney conditions | Chronic kidney disease | 12 (2.2) | 3 (1.3) | 9 (3.0) |

<sup>1</sup>Preexisting conditions were classified using CDC guidance on underlying medical conditions associated with higher risk of COVID-19 found at https://www.cdc.gov/coronavirus/2019-ncov/need-extra-precautions/people-with-medical-conditions.html.

**Table S3:** Estimated two-dose VE with each month following second dose receipt.

| Days after full vaccination | Estimated VE (95% CI) |
| --- | --- |
| 0 | 91.3 (83.8,95.4) |
| 30 | 86.4 (79.3,91.1) |
| 60 | 81.5 (74.7,86.6) |
| 90 | 76.4 (68.6,82.4) |
| 120 | 70.8 (60.4,78.6) |
| 150 | 64.7 (49.6,75.4) |
| 180 | 58.0 (36.1,72.4) |
| 210 | 50.8 (19.7,69.8) |
| 240 | 42.9 (-0.1,67.1) |
| 270 | 38.4 (-11.9,65.6) |

**Table S4:** Model comparisons for inter-dose interval and waning effects of two-dose vaccination.

| Model | BIC | AIC |
| --- | --- | --- |
| Inter-dose models |  |  |
| clogit(case~FV*DaysBetweenDoses + strata(age-sex-region-testDate) | 1097.038 | 1082.507 |
| clogit(case~ FV*daysPostDose2*recDosing2lev + strata(age-sex-region-testDate) | 1119.559 | 1085.652 |
| clogit(case~FV*daysPostDose2 + FV*DaysBetweenDoses + strata(age-sex-region-testDate) | 1104.798 | 1080.579 |
| Waning effect over time |  |  |
| clogit(case~FV*daysPostDose2 + strata(age-sex-region-test)) |  |  |
| Log transformation of days after participants were fully vaccinated | Did not converge | Did not converge |
| Linear transformation of days after participants were fully vaccinated | 1149.536 | 1130.153 |
| Square root transformation of days after participants were fully vaccinated | 1140.875 | 1126.338 |

Models are specified in R syntax for regression components. AIC: Akaike information criterion; BIC: Bayesian information criterion.

**Table S5:** Depletion-of-susceptible bias-adjusted VE estimates.

| Proportion uninfected at baseline |  | Bias-corrected VE as of December, 2021, by month when individuals were considered fully vaccinated with two doses, % (95% CI) |  |  |  |  |  |  |  |  |
| --- | --- | --- | --- | --- | --- | --- | --- | --- | --- | --- |
| Among the vaccinated | Among the unvaccinated | November | October | September | August | July | June | May | April | March |
| 100% | 100% | 86.5 (79.5, 91.1) | 81.7 (74.9, 86.7) | 76.6 (68.9, 82.6) | 71.1 (60.8, 78.9) | 65.2 (50.3, 75.7) | 59.3 (38.0, 73.2) | 53.2 (23.6, 71.2) | 46.4 (5.9, 69.1) | 38.9 (-14.5, 66.9) |
|  | 90% | 87.8 (81.5, 92.0) | 83.5 (77.4, 88.1) | 78.9 (72.0, 84.3) | 74.0 (64.7, 81.0) | 68.7 (55.3, 78.1) | 63.4 (44.2, 75.9) | 57.9 (31.3, 74.1) | 51.7 (15.3, 72.1) | 45.0 (-3.0, 70.2) |
|  | 80% | 89.2 (83.6, 92.9) | 85.4 (79.9, 89.4) | 81.3 (75.1, 86.1) | 76.9 (68.7, 83.1) | 72.2 (60.3, 80.6) | 67.4 (50.4, 78.6) | 62.5 (38.9, 77.0) | 57.1 (24.8, 75.2) | 51.1 (8.4, 73.6) |
| 90% | 100% | 85.0 (77.2, 90.1) | 79.7 (72.1, 85.3) | 74.0 (65.5, 80.6) | 67.9 (56.5, 76.5) | 61.3 (44.8, 73.0) | 54.8 (31.1, 70.3) | 48.0 (15.1, 68.0) | 40.4 (-4.5, 65.6) | 32.1 (-27.2, 63.3) |
|  | 90% | 86.5 (79.5, 91.1) | 81.7 (74.9, 86.7) | 76.6 (68.9, 82.6) | 71.1 (60.8, 78.9) | 65.2 (50.3, 75.7) | 59.3 (38.0, 73.2) | 53.2 (23.6, 71.2) | 46.4 (5.9, 69.1) | 38.9 (-14.5, 66.9) |
|  | 80% | 88.0 (81.7, 92.1) | 83.7 (77.7, 88.2) | 79.2 (72.4, 84.5) | 74.3 (65.2, 81.2) | 69.1 (55.8, 78.4) | 63.8 (44.8, 76.2) | 58.4 (32.1, 74.4) | 52.3 (16.4, 72.5) | 45.7 (-1.7, 70.6) |
| 80% | 100% | 83.1 (74.3, 88.9) | 77.1 (68.6, 83.4) | 70.8 (61.2, 78.2) | 63.9 (51.0, 73.6) | 56.5 (37.9, 69.7) | 49.1 (22.4, 66.6) | 41.5 (4.5, 64.1) | 32.9 (-17.6, 61.3) | 23.6 (-43.1, 58.7) |
|  | 90% | 84.8 (76.9, 90.0) | 79.4 (71.8, 85.1) | 73.7 (65.0, 80.4) | 67.5 (55.9, 76.2) | 60.9 (44.1, 72.7) | 54.2 (30.2, 69.9) | 47.3 (14.1, 67.6) | 39.6 (-5.8, 65.2) | 31.2 (-28.8, 62.8) |
|  | 80% | 86.5 (79.5, 91.1) | 81.7 (74.9, 86.7) | 76.6 (68.9, 82.6) | 71.1 (60.8, 78.9) | 65.2 (50.3, 75.7) | 59.3 (38.0, 73.2) | 53.2 (23.6, 71.2) | 46.4 (5.9, 69.1) | 38.9 (-14.5, 66.9) |

**Table S6:** Characteristics of participants who received a third dose ≥7 days before SARS-CoV-2 testing.

|  | All third dose recipients | Cases | Controls |
| --- | --- | --- | --- |
|  | *n* (%) | *n* (%) | *n* (%) |
|  | *N*=25 | *N*=3 | *N*=22 |
| Age (years) |  |  |  |
| 18-29 | 4 (16.0) | 0 (0.0) | 4 (18.2) |
| 30-49 | 8 (32.0) | 1 (33.3) | 7 (31.8) |
| 50-64 | 5 (20.0) | 1 (33.3) | 4 (18.2) |
| 65+ | 8 (32.0) | 1 (33.3) | 7 (31.8) |
| Sex |  |  |  |
| Male | 8 (32.0) | 0 (0.0) | 8 (36.4) |
| Female | 17 (68.0) | 3 (100.0) | 14 (63.6) |
| Month of receipt of third dose |  |  |  |
| August | 2 (8.0) | 0 (0.0) | 2 (9.1) |
| September | 5 (20.0) | 0 (0.0) | 5 (22.7) |
| October | 13 (52.0) | 2 (66.7) | 11 (50.0) |
| November | 5 (20.0) | 1 (33.3) | 4 (18.2) |
| Vaccine series |  |  |  |
| 3 doses BNT162b2 | 15 (60.0) | 2 (66.7) | 13 (59.1) |
| 3 doses mRNA-1273 | 7 (28.0) | 0 (0.0) | 7 (31.8) |
| 2 doses BNT162b2 + mRNA-1273 | 1 (4.0) | 0 (0.0) | 1 (4.5) |
| 2 doses mRNA-1273 + BNT162b2 | 2 (8.0) | 1 (33.3) | 1 (4.5) |
| Presence of preexisting conditions<sup>1</sup> |  |  |  |
| No conditions | 11 (44.0) | 2 (66.7) | 9 (40.9) |
| Any conditions | 14 (56.0) | 1 (33.3) | 13 (60.1) |

<sup>1</sup>Pre-existing conditions reported within the sample are enumerated in **Table S2**.

**Figure S1:**
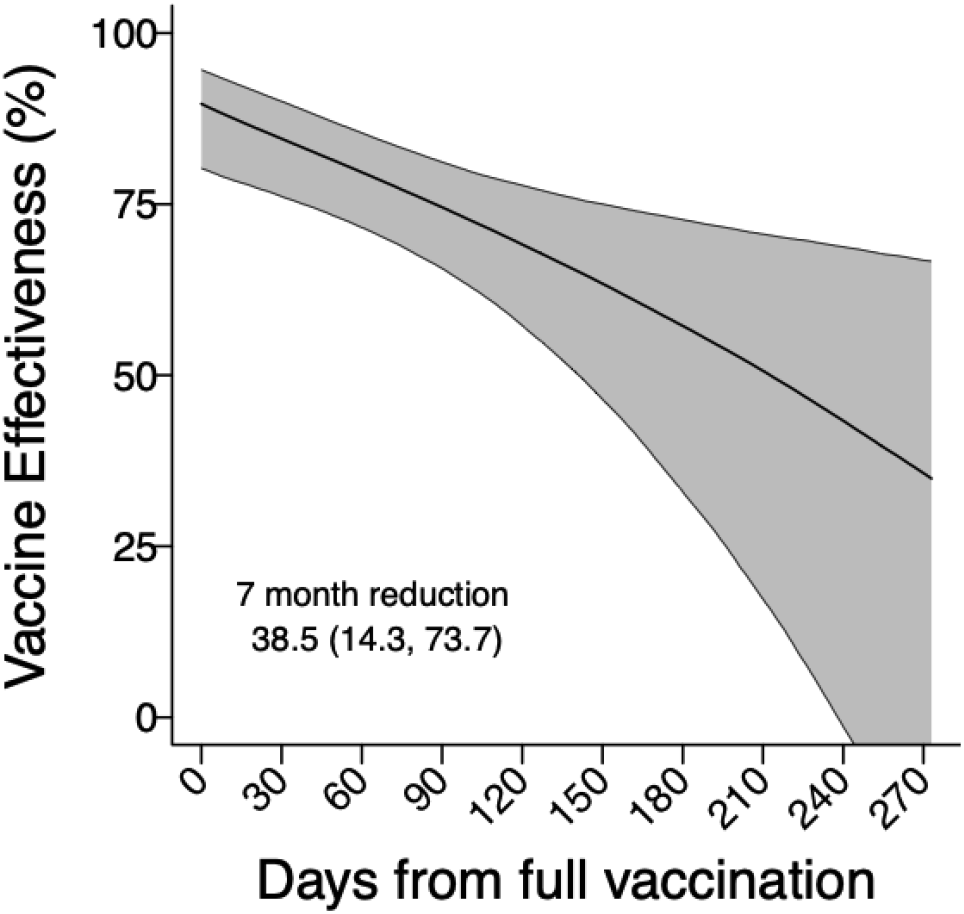
Two-dose vaccine effectiveness against symptomatic SARS-CoV-2 infection over time, excluding individuals who did not reference their vaccination card (*N*=114) at the time of their interview. Estimates are obtained in the same framework as those presented in **Figure 2** but excluding individuals who did not refer to their vaccination card (or other record of dates of receipt of each vaccine dose) during their interview.

**Figure S2:**
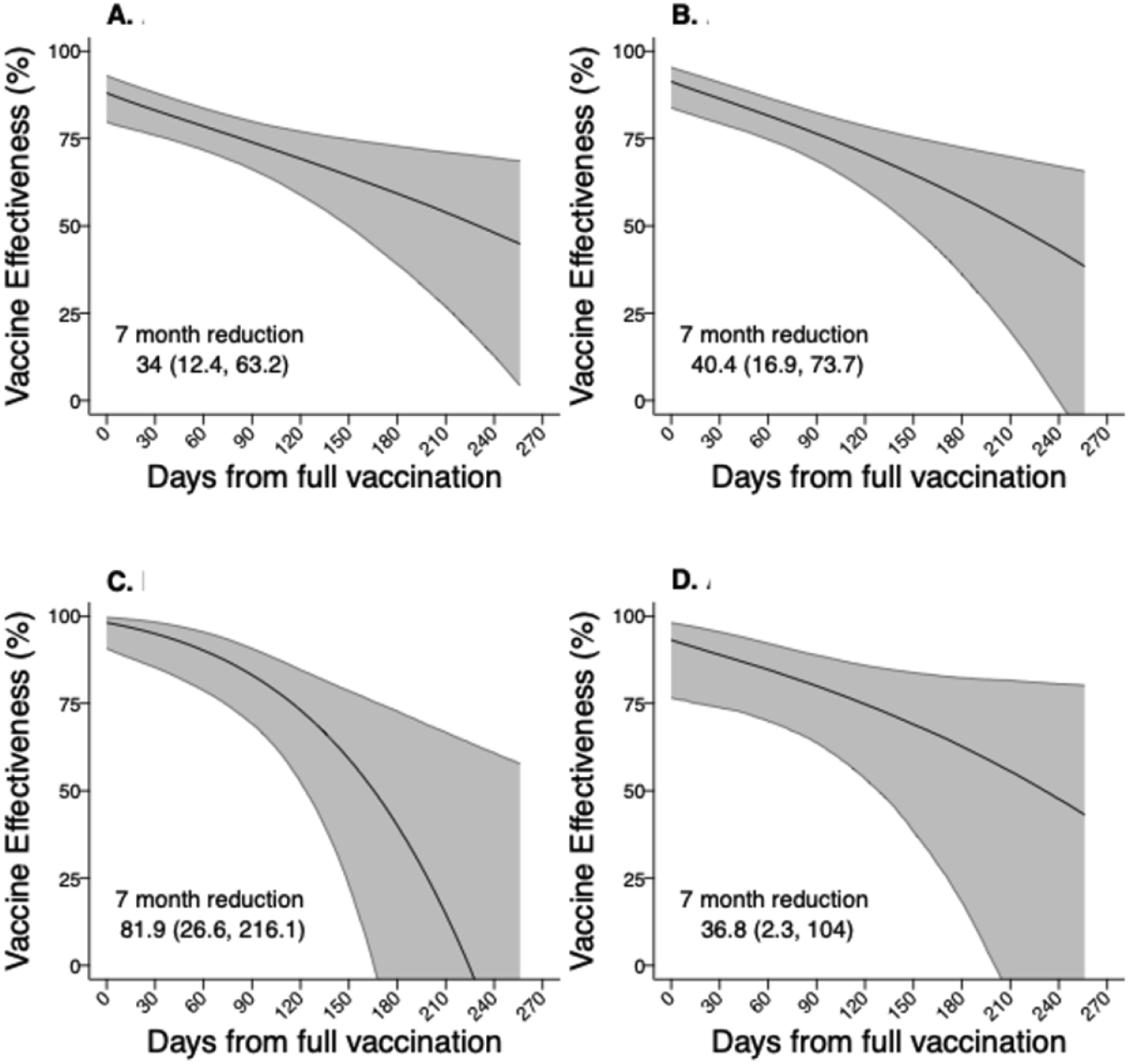
Two-dose vaccine effectiveness against alternative SARS-CoV-2 infection endpoints over time. We present estimates (obtained in the same framework as those presented in **Figure 2**) of two-dose vaccine effectiveness by time since individuals were considered fully vaccinated with two doses, for differing endpoint definitions: any infection (1300 cases with 1174 controls; **A**), SARS-CoV-2 infection with ≥1 symptom reported (*N*=1049 cases with 202 test-negative controls reporting symptoms; **B**), SARS-CoV-2 infection with fever and at least one respiratory symptom (*N*=205 cases with 18 test-negative controls reporting these symptoms; **C**), and SARS-CoV-2 infection for which participants received care in any setting (*N*=295 cases with 97 test-negative controls who also reported receiving care beyond testing; **D**).

**Figure S3:**
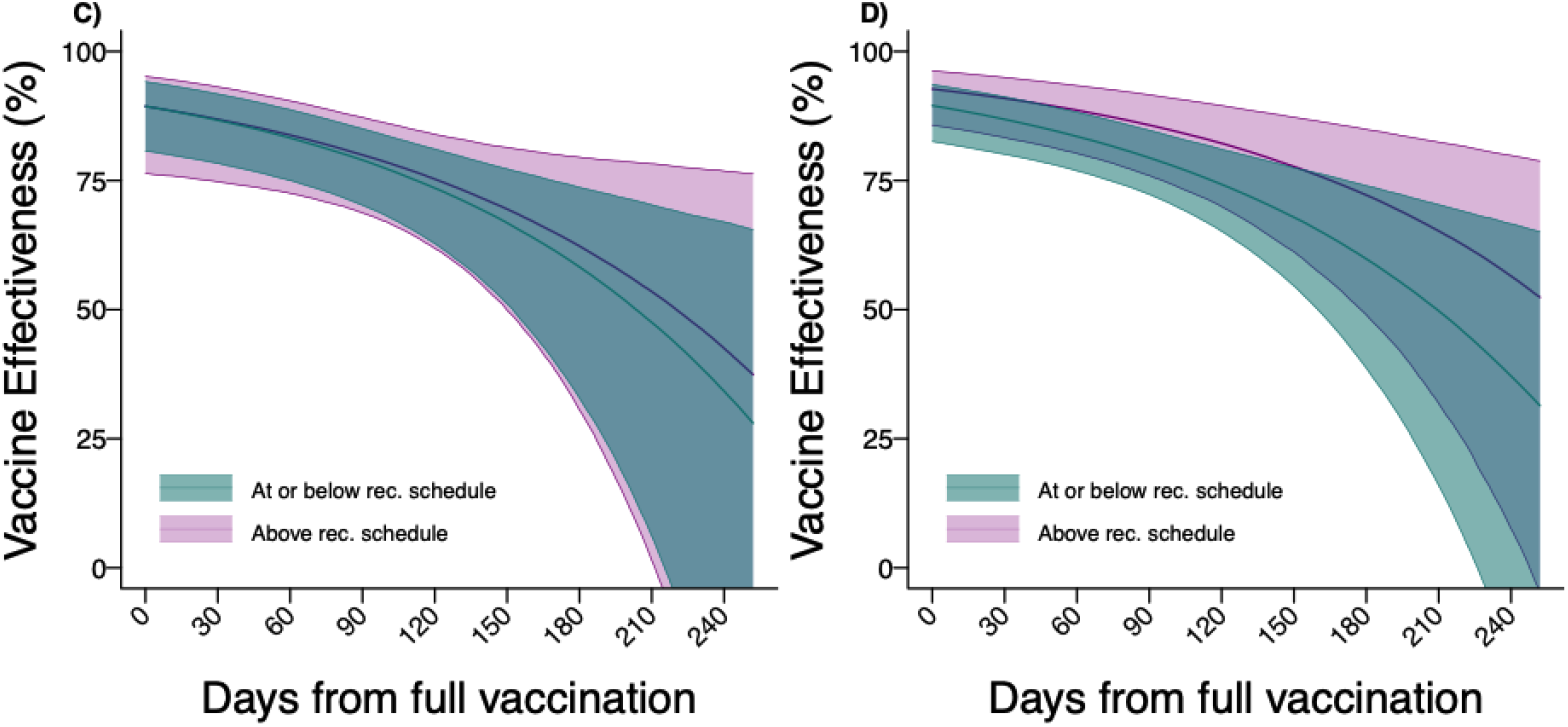
Long-term effectiveness of two vaccine doses against symptomatic SARS-CoV-2 infection by interval between receipt of first and second doses. We plot long-term estimates of VE, stratified for participants who received first and second doses spaced at or below the recommended intervals (≤21 days for BNT162b2 or ≤28 days for mRNA-1273) versus at longer intervals (>21 days for BNT162b2 or >28 days for mRNA-1273), estimated via conditional logistic regression models matching cases and controls on age group, sex, region, and week of SARS-CoV-2 test. Panels illustrate results of models assuming differing initial vaccine effectiveness between the groups only (**C**) or differences in both initial vaccine effectiveness and subsequent rates of waning (**D**). Model comparisons based on AIC and BIC are presented in **Table S4**.

**Figure S4:**
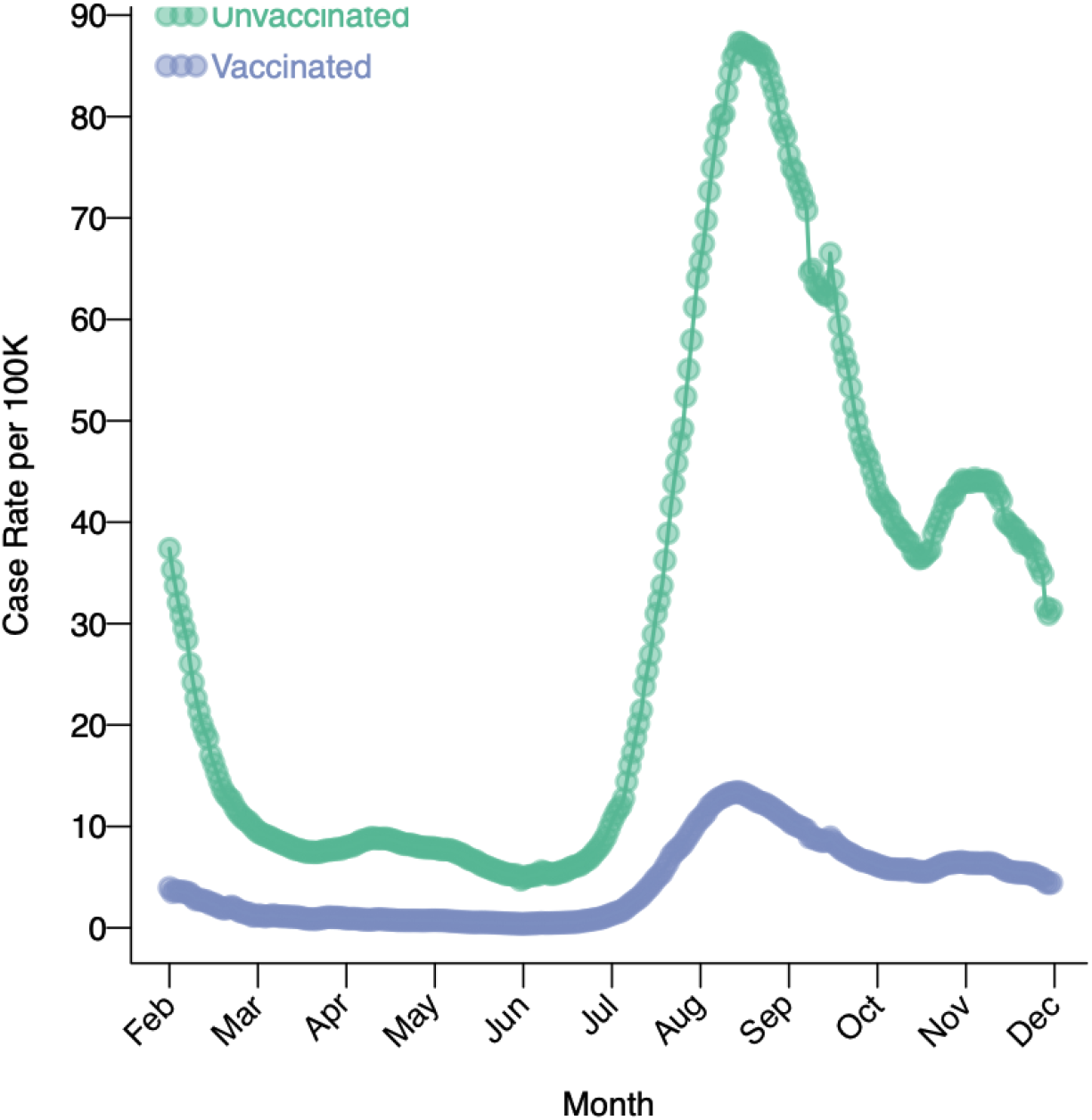
Incidence rates of daily reported SARS-CoV-2 infection in California, stratified by vaccination status (unvaccinated versus fully vaccinated with ≥2 doses), from February, 2021 to December, 2021. Incidence rates (presented as a seven-day moving average) were obtained through the publicly available California Health and Human Services Open Data Portal at [https://data.chhs.ca.gov/dataset/covid-19-post-vaccination-infection-data].

## REFERENCES

1. Sandmann FG, Jit M. Rapid COVID-19 vaccine rollout: immense success but challenges ahead. The Lancet Infectious Diseases [electronic article]. 2021;0(0). (https://www.thelancet.com/journals/laninf/article/PIIS1473-3099(21)00616-2/fulltext). (Accessed December 1, 2021)

2. Polack FP, Thomas SJ, Kitchin N, et al. Safety and Efficacy of the BNT162b2 mRNA Covid-19 Vaccine. New England Journal of Medicine. 2020;383(27):2603–2615.

3. Baden LR, El Sahly HM, Essink B, et al. Efficacy and Safety of the mRNA-1273 SARS-CoV-2 Vaccine. New England Journal of Medicine. 2021;384(5):403–416.

4. Sadoff J, Le Gars M, Shukarev G, et al. Interim Results of a Phase 1–2a Trial of Ad26.COV2.S Covid-19 Vaccine. N Engl J Med. 2021;NEJMoa2034201.

5. COVIDVaxView | CDC. 2021;(https://www.cdc.gov/vaccines/imz-managers/coverage/covidvaxview/index.html). (Accessed November 30, 2021)

6. Brosh-Nissimov T, Orenbuch-Harroch E, Chowers M, et al. BNT162b2 vaccine breakthrough: clinical characteristics of 152 fully vaccinated hospitalized COVID-19 patients in Israel. Clinical Microbiology and Infection. 2021;27(11):1652–1657.

7. Tenforde MW, Patel MM, Ginde AA, et al. Effectiveness of Severe Acute Respiratory Syndrome Coronavirus 2 Messenger RNA Vaccines for Preventing Coronavirus Disease 2019 Hospitalizations in the United States. Clinical Infectious Diseases. 2021;ciab687.

8. CDC. Quarantine & Isolation. Centers for Disease Control and Prevention. 2022; (https://www.cdc.gov/coronavirus/2019-ncov/your-health/quarantine-isolation.html). (Accessed June 3, 2022)

9. Saad-Roy CM, Wagner CE, Baker RE, et al. Immune life history, vaccination, and the dynamics of SARS-CoV-2 over the next 5 years. Science. 2020;370(6518):811–818.

10. Tartof SY, Slezak JM, Fischer H, et al. Effectiveness of mRNA BNT162b2 COVID-19 vaccine up to 6 months in a large integrated health system in the USA: a retrospective cohort study. The Lancet. 2021;398(10309):1407–1416.

11. Goldberg Y, Mandel M, Bar-On YM, et al. Waning immunity of the BNT162b2 vaccine: A nationwide study from Israel. 2021 (Accessed October 31, 2021).(https://www.medrxiv.org/content/10.1101/2021.08.24.21262423v1). (Accessed October 31, 2021)

12. Chemaitelly H, Tang P, Hasan MR, et al. Waning of BNT162b2 Vaccine Protection against SARS-CoV-2 Infection in Qatar. New England Journal of Medicine. 2021;0(0):null.

13. Lopez Bernal J, Andrews N, Gower C, et al. Effectiveness of Covid-19 Vaccines against the B.1.617.2 (Delta) Variant. New England Journal of Medicine. 2021;0(0):null.

14. Seppälä E, Veneti L, Starrfelt J, et al. Vaccine effectiveness against infection with the Delta (B.1.617.2) variant, Norway, April to August 2021. Eurosurveillance. 2021;26(35):2100793.

15. Nasreen S, Chung H, He S, et al. Effectiveness of mRNA and ChAdOx1 COVID-19 vaccines against symptomatic SARS-CoV-2 infection and severe outcomes with variants of concern in Ontario. 2021 (Accessed November 1, 2021).(https://www.medrxiv.org/content/10.1101/2021.06.28.21259420v3). (Accessed November 1, 2021)

16. Bruxvoort KJ, Sy LS, Qian L, et al. Effectiveness of mRNA-1273 against Delta, Mu, and other emerging variants. 2021 (Accessed November 30, 2021).(https://www.medrxiv.org/content/10.1101/2021.09.29.21264199v1). (Accessed November 30, 2021)

17. Scott L, Hsiao N, Moyo S, et al. Track Omicron’s spread with molecular data. Science [electronic article]. 2021;(https://www.science.org/doi/abs/10.1126/science.abn4543). (Accessed January 21, 2022)

18. Pulliam JRC, Schalkwyk C van, Govender N, et al. Increased risk of SARS-CoV-2 reinfection associated with emergence of the Omicron variant in South Africa. 2021 (Accessed January 21, 2022).(https://www.medrxiv.org/content/10.1101/2021.11.11.21266068v2). (Accessed January 21, 2022)

19. Lewnard JA, Hong VX, Patel MM, et al. Clinical outcomes among patients infected with Omicron (B.1.1.529) SARS-CoV-2 variant in southern California. 2022 (Accessed January 21, 2022).(https://www.medrxiv.org/content/10.1101/2022.01.11.22269045v1). (Accessed January 21, 2022)

20. Smith P, Rodrigues L, Fine P. Assessment of the Protective Efficacy of Vaccines against Common Diseases Using Case-Control and Cohort Studies. International Journal of Epidemiology. 1984;13(1):87–93.

21. Lipsitch M. Challenges of Vaccine Effectiveness and Waning Studies. Clin Infect Dis. 2019;68(10):1631–1633.

22. Lewnard JA, Tedijanto C, Cowling BJ, et al. Measurement of Vaccine Direct Effects Under the Test-Negative Design. Am J Epidemiol. 2018;187(12):2686–2697.

23. León TM. COVID-19 Cases and Hospitalizations by COVID-19 Vaccination Status and Previous COVID-19 Diagnosis — California and New York, May–November 2021. MMWR Morb Mortal Wkly Rep [electronic article]. 2022;71. (https://www.cdc.gov/mmwr/volumes/71/wr/mm7104e1.htm). (Accessed February 7, 2022)

24. Yola M, Lucien A. Evidence of the depletion of susceptibles effect in non-experimental pharmacoepidemiologic research. Journal of Clinical Epidemiology. 1994;47(7):731–737.

25. Kahn R, Schrag SJ, Verani JR, et al. Identifying and Alleviating Bias Due to Differential Depletion of Susceptible People in Postmarketing Evaluations of COVID-19 Vaccines. American Journal of Epidemiology. 2022;kwac015.

26. Self WH. Comparative Effectiveness of Moderna, Pfizer-BioNTech, and Janssen (Johnson & Johnson) Vaccines in Preventing COVID-19 Hospitalizations Among Adults Without Immunocompromising Conditions — United States, March–August 2021. MMWR Morb Mortal Wkly Rep [electronic article]. 2021;70. (https://www.cdc.gov/mmwr/volumes/70/wr/mm7038e1.htm). (Accessed November 30, 2021)

27. Iacobucci G, Mahase E. Covid-19 vaccination: What’s the evidence for extending the dosing interval? BMJ. 2021;372:n18.

28. California S of. Vaccination progress data. (https://covid19.ca.gov/vaccination-progress-data/). (Accessed October 19, 2021)

29. Andrejko KL, Pry J, Myers JF, et al. Prevention of Coronavirus Disease 2019 (COVID-19) by mRNA-Based Vaccines Within the General Population of California. Clinical Infectious Diseases. 2021;ciab640.

30. Andrejko KL, Pry J, Myers JF, et al. Predictors of Severe Acute Respiratory Syndrome Coronavirus 2 Infection Following High-Risk Exposure. Clinical Infectious Diseases. 2021;ciab1040.

31. Andrejko KL, Pry JM, Myers JF, et al. Effectiveness of Face Mask or Respirator Use in Indoor Public Settings for Prevention of SARS-CoV-2 Infection — California, February–December 2021. MMWR Morb. Mortal. Wkly. Rep. 2022;71(6):212–216.

32. Lewnard J, Hong V, Patel M, et al. Clinical outcomes associated with Omicron (B.1.1.529) variant and BA.1/BA.1.1 or BA.2 subvariant infection in southern California. Nature Medicine. 2022;

33. Tartof SY, Slezak JM, Puzniak L, et al. Durability of BNT162b2 vaccine against hospital and emergency department admissions due to the omicron and delta variants in a large health system in the USA: a test-negative case–control study. The Lancet Respiratory Medicine. 2022;S2213260022001011.

34. CDC. People with Certain Medical Conditions. Centers for Disease Control and Prevention. 2020;(https://www.cdc.gov/coronavirus/2019-ncov/need-extra-precautions/people-with-medical-conditions.html). (Accessed November 9, 2021)

35. Lewnard JA, Patel MM, Jewell NP, et al. Theoretical Framework for Retrospective Studies of the Effectiveness of SARS-CoV-2 Vaccines. Epidemiology. 2021;32(4):508–517.

36. Dean NE, Hogan JW, Schnitzer ME. Covid-19 Vaccine Effectiveness and the Test-Negative Design. New England Journal of Medicine. 2021;385(15):1431–1433.

37. Payne RP, Longet S, Austin JA, et al. Immunogenicity of standard and extended dosing intervals of BNT162b2 mRNA vaccine. Cell. 2021;184(23):5699-5714.e11.

38. Wu SL, Mertens AN, Crider YS, et al. Substantial underestimation of SARS-CoV-2 infection in the United States. Nat Commun. 2020;11(1):4507.

39. Oran DP, Topol EJ. The Proportion of SARS-CoV-2 Infections That Are Asymptomatic. Ann Intern Med. 2021;174(5):655–662.

40. Shenai MB, Rahme R, Noorchashm H. Equivalency of Protection from Natural Immunity in COVID-19 Recovered Versus Fully Vaccinated Persons: A Systematic Review and Pooled Analysis. 2021 (Accessed November 9, 2021).(https://www.medrxiv.org/content/10.1101/2021.09.12.21263461v1). (Accessed November 9, 2021)

41. COVID-19 Post-Vaccination Infection Data - California Health and Human Services Open Data Portal. (https://data.chhs.ca.gov/dataset/covid-19-post-vaccination-infection-data). (Accessed October 28, 2021)

42. Mehrotra ML, Lim E, Lamba K, et al. CalScope: Monitoring SARS-CoV-2 Seroprevalence from Vaccination and Prior Infection in Adults and Children in California May 2021 to July 2021. 2021 (Accessed December 13, 2021).(https://www.medrxiv.org/content/10.1101/2021.12.09.21267565v1). (Accessed December 13, 2021)

43. Lamba K, Bradley H, Shioda K, et al. SARS-CoV-2 Cumulative Incidence and Period Seroprevalence: Results From a Statewide Population-Based Serosurvey in California. Open Forum Infect Dis. 2021;8(8):ofab379.

44. Israel A, Merzon E, Schäffer AA, et al. Elapsed time since BNT162b2 vaccine and risk of SARS-CoV-2 infection in a large cohort. Infectious Diseases (except HIV/AIDS); 2021 (Accessed October 19, 2021).(http://medrxiv.org/lookup/doi/10.1101/2021.08.03.21261496). (Accessed October 19, 2021)

45. Mizrahi B, Lotan R, Kalkstein N, et al. Correlation of SARS-CoV-2 Breakthrough Infections to Time-from-vaccine; Preliminary Study. Epidemiology; 2021 (Accessed October 19, 2021).(http://medrxiv.org/lookup/doi/10.1101/2021.07.29.21261317). (Accessed October 19, 2021)

46. Andrews N, Tessier E, Stowe J, et al. Vaccine effectiveness and duration of protection of Comirnaty, Vaxzevria and Spikevax against mild and severe COVID-19 in the UK. Epidemiology; 2021 (Accessed October 19, 2021).(http://medrxiv.org/lookup/doi/10.1101/2021.09.15.21263583). (Accessed October 19, 2021)

47. Cox LS, Bellantuono I, Lord JM, et al. Tackling immunosenescence to improve COVID-19 outcomes and vaccine response in older adults. The Lancet Healthy Longevity. 2020;1(2):e55–e57.

48. Anand S, Montez-Rath ME, Han J, et al. SARS-CoV-2 Vaccine Antibody Response and Breakthrough Infection in Patients Receiving Dialysis. Ann Intern Med [electronic article]. 2021;(https://www.acpjournals.org/doi/10.7326/M21-4176). (Accessed February 7, 2022)

49. Centers for Disease Control and Prevention. CDC Strengthens Recommendations and Expands Eligibility for COVID-19 Booster Shots. Centers for Disease Control and Prevention. 2022;(https://www.cdc.gov/media/releases/2022/s0519-covid-booster-acip.html). (Accessed June 3, 2022)

50. Amirthalingam G, Bernal JL, Andrews NJ, et al. Higher serological responses and increased vaccine effectiveness demonstrate the value of extended vaccine schedules in combatting COVID-19 in England. 2021 (Accessed November 1, 2021).(https://www.medrxiv.org/content/10.1101/2021.07.26.21261140v1). (Accessed November 1, 2021)

51. Ferdinands JM. Waning 2-Dose and 3-Dose Effectiveness of mRNA Vaccines Against COVID-19–Associated Emergency Department and Urgent Care Encounters and Hospitalizations Among Adults During Periods of Delta and Omicron Variant Predominance — VISION Network, 10 States, August 2021–January 2022. MMWR Morb Mortal Wkly Rep [electronic article]. 2022;71. (https://www.cdc.gov/mmwr/volumes/71/wr/mm7107e2.htm). (Accessed February 11, 2022)

